# Multiple imputation assuming missing at random: auxiliary imputation variables that only predict missingness can increase bias due to data missing not at random

**DOI:** 10.1101/2023.10.17.23297137

**Authors:** Elinor Curnow, Rosie P Cornish, Jon E Heron, James R Carpenter, Kate Tilling

## Abstract

Epidemiological studies often have missing data, which are commonly handled by multiple imputation (MI). MI is valid (given correctly-specified models) if data are missing at random, conditional on the observed data, but not (unless additional information is available) if data are missing not at random (MNAR). In this paper we explore a previously-suggested strategy, namely, including an auxiliary variable predictive of missingness but not the missing data in the imputation model, when data are MNAR. We quantify, algebraically and by simulation, the magnitude of additional bias of the MI estimator, over and above any bias due to data MNAR, from including such an auxiliary variable. We demonstrate that where missingness is caused by the outcome, additional bias can be substantial when the outcome is partially observed. Furthermore, if missingness is caused by the outcome and the exposure, additional bias can be even larger, when either the outcome or exposure is partially observed. When using MI, it is important to identify, through a combination of data exploration and considering plausible casual diagrams and missingness mechanisms, the auxiliary variables most predictive of the missing data (in addition to all variables required for the analysis model and/or to minimise bias due to MNAR).

## 1. Introduction

Epidemiological studies often have missing data, with multiple imputation (MI) a commonly-used, flexible, and general method for analysing partially observed datasets [1]. When imputation models are appropriately specified and compatible with the substantive analysis model (*i.e*. containing the same variables and including any required non-linear terms or interactions) [2], MI gives valid inferences if data are missing completely at random (MCAR) or missing at random (MAR), conditional on the observed data, but not (unless additional information is available) if data are missing not at random (MNAR) (Table 1).

**Table 1.** Missing data definitions.

| Term | Definition |
| --- | --- |
| Complete Records Analysis (CRA) | Analysis is restricted to subjects who have complete data for all variables in the analysis model. |
| Missing Completely At Random (MCAR) | The probability that data are missing is independent of the observed and missing values of variables in the analysis model, and of any related variables. Data can be MCAR if missingness is caused by a variable independent of those in the analysis model *e.g.* if missingness is for administrative reasons. |
| Missing At Random (MAR) | Given the observed data, the probability that data are missing is independent of the true values of the partially observed variable. Any systematic differences between the observed and missing values can be explained by associations with the observed data. |
| Missing Not At Random (MNAR) | If data are not MCAR nor MAR, data are said to be MNAR. The probability that data are missing depends on the (unobserved) values of the partially observed variable, even after conditioning on the observed data. |
| Multiple Imputation (MI) | MI is a method for handling missing data. It consists of three steps:<br>1. An imputation model is fitted to the observed data (this is usually some form of regression model). The missing values are replaced with draws (“imputed”) from its predictive distribution (after first perturbing the model parameters). This imputation stage is carried out multiple (M) times, to give M completed datasets.<br>2. The analysis model is fitted to each of the M completed datasets.<br>3. The M sets of results are combined using Rubin’s rules, [3] to correctly account for the uncertainty about the missing values. |
| Predictive Mean Matching (PMM) | PMM is an MI approach that uses an alternative method in Step 1 of the MI process: instead of imputing missing values directly from the conditional predictive distribution of the missing data given the observed data, each missing value is replaced with an observed value randomly chosen from a donor pool anchored on the conditional predicted mean. |
| Auxiliary variable | A variable that is not in the analysis model but that is included as a predictor in the imputation model. |

For example, consider a longitudinal cohort study where we are interested in the association between a partially observed outcome, child’s IQ, and a fully observed exposure, duration of breastfeeding. If the probability that child’s IQ is missing (its “missingness”) is related to neither observed nor missing values of child’s IQ, given the observed data for the other analysis model variables, and all these variables are included in both analysis and imputation models, then both MI and complete records analysis (CRA) estimates of the outcome-exposure association will be unbiased [4]. On the other hand, suppose that missingness in child’s IQ is caused by child’s IQ itself, as depicted in the causal diagram (or directed acyclic graph, DAG) in Figure 1 (with *X, Y*, and *R*_*ind*_ denoting our exposure (duration of breastfeeding), outcome (child’s IQ), and missingness indicator, respectively). In this case, even after conditioning on the observed data, child’s IQ is MNAR. Since child’s IQ is the outcome of the substantive analysis, both CRA and MI estimates of the outcome-exposure association will be biased [4].

**Figure 1.**
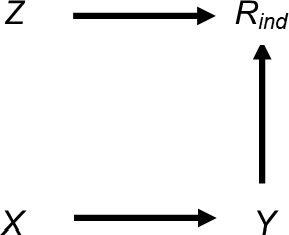
Directed acyclic graph depicting the relationship between outcome Y, exposure X, missingness indicator R_ind_, and potential auxiliary variable Z. Lines indicate related variables, with arrows indicating the direction of the relationship; absent lines represent variables with no direct causal relation

Common strategies when data are suspected to be MNAR include exploring the sensitivity of MI results to departures from the MAR assumption using a “pattern mixture” approach [5], which allows the observed and missing values to differ by a value, or set of values, δ (the “sensitivity parameter”); applying an MI method that can accommodate data MNAR [6]; or including a “proxy” for the partially observed variable (*i.e*. a variable that is predictive of the missing values) as an auxiliary variable (see Table 1) in the imputation model [7]. In this paper, we highlight the consequences of using an inclusive strategy as has been suggested previously (and, anecdotally, is common practice) [8-10]: including a predictor of missingness as an auxiliary variable in the imputation model. We demonstrate that when data are MNAR and such a variable is, in truth, unrelated to the partially observed variable, then the bias due to data being MNAR may be increased rather than reduced, in a similar way to bias amplification in the presence of unmeasured confounding when conditioning on variables that only influence an exposure [11]. Returning to our example, including the proxy “child’s educational attainment score” in the imputation model for child’s IQ may reduce the bias in the exposure-outcome association due to child’s IQ being MNAR [7, 12] because child’s educational attainment score is highly correlated with child’s IQ. However, including a predictor of missingness in the imputation model where this is unrelated to child’s IQ (denoted by *Z* in Figure 1, *e.g*. whether the mother smoked during pregnancy) may increase the bias of the MI estimate. Note (depending on the magnitude and direction of the associations), bias due to data MNAR may also increase even if auxiliary variables are predictive of both the missing values and missingness, particularly if the auxiliary variable is a collider [13].

In this paper we quantify the magnitude of the additional bias of the MI estimator due to using an auxiliary variable that predicts missingness, but not the values of the partially observed variable itself, when data are MNAR. By “additional bias”, we mean the difference between the MI estimator when including a predictor of missingness in the imputation model (as well as the other analysis model variables) and the MI estimator when including just the other analysis model variables in the imputation model (noting that both estimators will yield biased estimates of the true outcome-exposure association when data are MNAR). We consider settings in which either the outcome or exposure are MNAR, where the partially observed variable is either continuous or binary, and where missingness is caused by the partially observed variable itself and/or another related variable. We quantify the additional bias using algebraic methods and by simulation, and illustrate our results using the real data example described above. Throughout the paper, we assume that MI is performed by replacing missing values with draws from a suitable regression model (*i.e*. a linear or logistic regression model when the partially observed variable is continuous or binary, respectively) using a linear combination of the specified predictors. We focus on this approach, rather than *e.g*. predictive mean matching (PMM) [14] because MI using draws from a correctly specified model will generally yield more precise estimates than PMM [15]. All analyses were conducted using Stata (17.0, StataCorp LLC, College Station, TX). Stata code to perform the simulation studies is included in Supplementary Material, Section S6. Stata code to perform the real data analysis is included in Supplementary Material, Section S7.

## 2. Scenario 1. Additional bias of the MI estimator from including a predictor only of missingness in the imputation model when continuous outcome *Y* is partially observed and missingness is caused by *Y*

### 2.1. Methods

We first consider the setting in Figure 1, discussed above, in more detail. This simplified setting is chosen to give insights into the more complex settings that typically occur in epidemiological practice. We are interested in the relationship between a continuous outcome *Y* and a continuous exposure *X*, with *β*_*YX*_ denoting the parameter of interest. We assume that *X* is fully observed and *Y* is partially observed, with variable *R*_*ind*_ denoting the missingness indicator for *Y* (*R*_*ind*_ = 1 if *Y* is observed, and 0 otherwise) and *π*_1_ denoting the probability that *R*_*ind*_ = 1, or *π*_1_ = P(*R*_*ind*_ = 1). Our substantive model is simply the regression of *Y* on *X*; we do not adjust for (fully observed) continuous variable *Z* because it does not confound the *X*-*Y* relationship. Since *Y* is MNAR, with missingness caused by *Y* itself, the MI estimator will be biased (as will CRA).

### 2.2. Maximum additional bias of the MI estimator

Here we provide general expressions for the maximum additional bias of the MI estimator (when using *X* and *Z* as predictors in the imputation model for *Y* compared with just *X*), when continuous outcome *Y* is MNAR and missingness is caused by *Y*. A full derivation of the results is included in the Supplementary Material (Section S1). Equations were verified by simulation (Supplementary Material Section S2).

We assume that *Y, X, Z*, and *R* are normally distributed, where *R* is a variable with mean *μ*_*R*_ and variance *V*_*R*_ such that 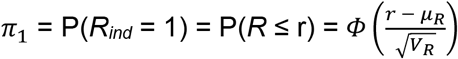, with *Φ* denoting the cumulative distribution function of the standard normal distribution. We assume that both *Y* and *R* are a linear combination of the variable(s) causing them plus an error term (with *X* and *Z* having no direct causes), with no interactions, all errors uncorrelated, no model mis-specification, and no measurement error. Finally, we assume an ordinary least squares (OLS) estimator is used to obtain estimates in both analysis and imputation models.

In this setting, the maximum additional bias of the MI estimator is equal to *β*_*YX*|*Z,R*_ − *β*_*YX*|*R*_. This follows from the argument of Curnow *et al*. [16], such that the MI estimator of *β*_*YX*_ (denoted by 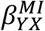) equals the regression parameter for *X* from the imputation model for *Y* based on records with observed values of *Y* (we denote this parameter by 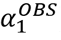). In other words, when the imputation model includes only *X* as a predictor, 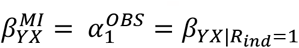 Curnow *et al*. showed that, in general, 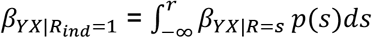 where *r* = *Φ*^−1^(*π*_1_) and *p*(*s*) denotes the probability that *R* = *s* given *s* ∈ (−∞, *r*]. Similarly, when the imputation model includes *X* and *Z* as predictors, 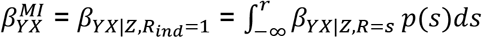. Thus it follows that the additional bias of the MI estimator from including *Z*, as well as *X*, in the imputation model for *Y* is equal to: 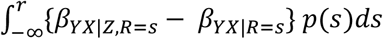. Furthermore, Curnow *et al*. showed that any bias in the MI estimator will increase with the proportion of missing values, and will tend to its maximum value as the proportion of missing values tends to one (or equivalently, since r = *Φ*^−1^(*π*_1_), as *r* tends to −∞). In this case, *p*(*s*) ≈ 0 for all *s* < *r*, with *p*(*r*) ≈ 1. Hence, the maximum additional bias is equal to: *β*_*YX*|*Z,R*=*r*_ − *β*_*YX*|*R*=*r*_, or more generally, *β*_*YX*|*Z,R*_ − *β*_*YX*|*R*_, since the magnitude of this expression does not depend on the value of *r* (see Supplementary Material, Section S1).

### 2.3. Maximum additional bias of the MI estimator in terms of the direct effect sizes

We next provide a general expression for the maximum additional bias of the MI estimator in terms of the direct effect sizes and error variances, such that:

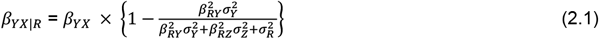

and

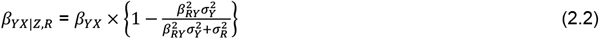

where the direct effect sizes are denoted by *β*_.._, *e.g. β*_*RY*_ denotes the direct effect of *Y* on *R*, and the error variances are denoted by *σ*^2^, *e.g. σ*^2^ denotes the error variance of *Y*.

Since 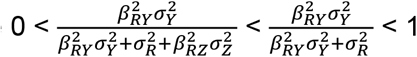 (assuming all parameters are non-zero), |*β*_*YX*|*Z,R*_| < |*β*_*YX*|*R*_| < |*β*_*YX*_|, that is, the MI estimator of *β*_*YX*_ will be biased towards zero, with the bias greater in magnitude when the imputation model includes *X* and *Z* as predictors than when it includes only *X*.

Then the maximum additional bias of the MI estimator from including *Z* as a predictor is:

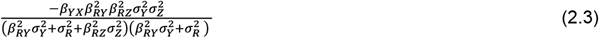

Equation (2.3) shows that the magnitude of the maximum additional bias will depend on the strength of the *Y*-*X, R*-*Y*, and *R*-*Z* relationships, as well as on the size of the error variances. There will be no additional bias if at least one of *β*_*YX*_, *β*_*RY*_, or *β*_*RZ*_ is equal to zero, consistent with the underlying DAG (Figure 1). Note that we can also express the effect on the MI estimator of including *Z* as a predictor in the imputation model in terms of bias amplification (defined as the bias of *β*_*YX*|*Z,R*_ divided by the bias of *β*_*YX*|*R*_): when *Z* (as well as *X*) is included in the imputation model for *Y*, the maximum bias due to *Y* being MNAR is amplified by a factor of:

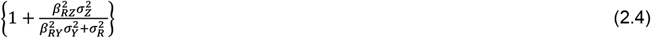

Note that if instead *X* was partially observed and *Y* was fully observed, MI would yield unbiased results (given a correctly specified imputation model) because in this case *R* would not be related to *X* after conditioning on *Y*. However, CRA would still be invalid because missingness depends on the analysis outcome.

### 2.4. Illustration of maximum additional bias of the MI estimator

We illustrate how the maximum additional bias varies with the direct effect sizes using a numerical example. In this example, we used moderate values of the direct effect sizes *β*_*YX*_, *β*_*RY*_, and *β*_*RZ*_ (relative to the error variances 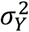 and 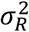, which were equal to one): direct effect sizes were each set to 0.00, 0.25, 0.50, 0.75, or 1.00. For *β*_*RY*_ and *β*_*RZ*_, note that these values correspond approximately to odds ratios (from a logistic regression model for *R*_*ind*_) of 1.00, 1.50, 2.30, 3.50, or 5.30 (using the general rule for transforming a parameter from a logistic to a probit model [17]). Figure 2 illustrates the impact of the direct effect sizes *β*_*YX*_, *β*_*RY*_, and *β*_*RZ*_ on various measures of bias. Panel A depicts the maximum bias of the MI estimator (due to *Y* being MNAR) when the imputation model includes only *X* as a predictor. Panels B-D depict the maximum additional bias (compared to the bias due to *Y* being MNAR), the maximum total bias (the sum of the maximum bias due to *Y* being MNAR and the maximum additional bias), and the maximum relative additional bias (maximum additional bias multiplied by 100, divided by *β*_*YX*_), respectively, when the imputation model includes both *X* and *Z* as predictors. The distribution of each box-plot is due to variation in *β*_*RY*_.

**Figure 2.**
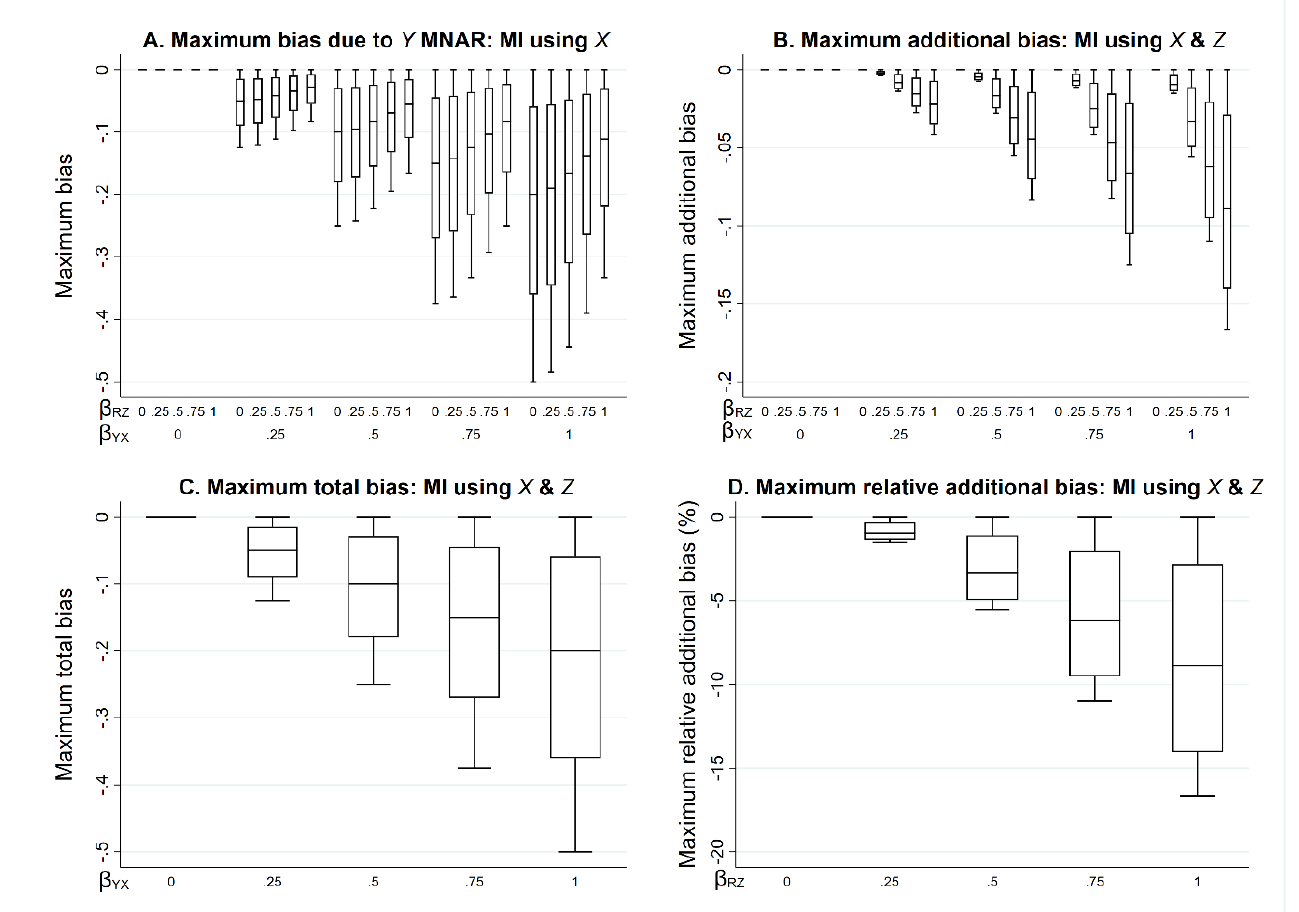
Bias of the MI estimator of β_YX_ when continuous outcome Y is missing not at random, with missingness caused by Y itself, and the imputation model includes exposure X, or X and a predictor of missingness but not the missing values, Z, varying the direct effect sizes β_YX_, β_RY_, and β_RZ_. Panel A depicts the maximum bias when the imputation model includes X. Panels B-D depict the maximum additional bias, maximum total bias, and maximum relative additional bias, respectively, when the imputation model includes X and Z. The distribution of each box-plot is due to variation in β_RY_. Note that maximum total depends on β_YX_ and β_RY_ but not β_RZ_; maximum relative additional bias depends on β_RZ_ and β_RY_ but not β_YX_.

Each measure of bias is equal to zero if *β*_*YX*_ is equal to zero (additionally, the maximum additional bias is equal to zero if any of the direct effect sizes are equal to zero), and negative otherwise. The maximum bias due to *Y* being MNAR increases in magnitude with *β*_*YX*_, but for a given value of *β*_*YX*_, decreases in magnitude as *β*_*RZ*_ increases. However, the maximum additional bias increases in magnitude with each of the direct effect sizes, as do the maximum total bias (which depends on *β*_*YX*_ and *β*_*RY*_ but not *β*_*RZ*_) and the maximum relative additional bias (which depends on *β*_*RZ*_ and *β*_*RY*_ but not *β*_*YX*_). Note that all parameters have a zero or positive value in this illustration. However, if, for example, we take the same parameter values as mentioned above for *β*_*RY*_ and *β*_*RZ*_, but set *β*_*YX*_ to negative values, then the measures of bias would be of the same magnitude but positive.

When the relationships are as depicted in Figure 1, but *Y* is binary, the results described here will still approximately apply (see Supplementary Material, Figure S1). This follows by assuming that *Y* has an underlying normal distribution (which is valid provided the probability of each value of *Y* is not close to 0 or 1).

## 3. Scenario 2. Additional bias of the MI estimator from including a predictor only of missingness in the imputation model when continuous outcome *Y* or continuous exposure *X* are partially observed and missingness is related to *Y* via an unmeasured variable

We next consider the setting in which missingness of the partially observed variable (either *Y* or *X*) is related to *Y* via an unmeasured variable, U, as depicted in Figure 3. In this setting (given the same assumptions and using the same analysis model and MI method as in the previous scenario), we would expect the CRA estimator and the MI estimator to be biased because missingness is related to our analysis outcome *Y* (conditional on *X*), via *U*. However, in the special case in which partially observed variable *Y* is continuous and the analysis model is a linear regression, both the CRA and MI estimators (using either *X*, or *X* and *Z*, as predictors in the imputation model for *Y*) are unbiased. Proof is provided in Supplementary Material, Section S3. Note that this is not the case if *Y* is binary, although the bias is generally small (see Supplementary Material, Figures S2-3).

**Figure 3.**
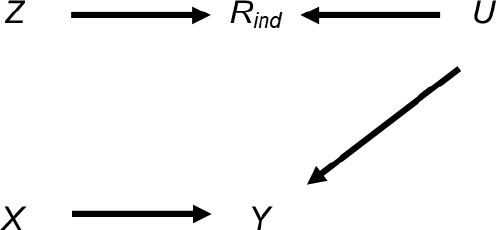
Directed acyclic graph depicting the relationship between outcome Y, exposure X, missingness indicator R_ind_, potential auxiliary variable Z, and unmeasured variable U. Lines indicate related variables, with arrows indicating the direction of the relationship; absent lines represent variables with no direct causal relation

When *X* is partially observed, the MI estimator (using either *Y*, or *Y* and *Z*, as predictors in the imputation model for *X*) will be biased because missingness is related to *X*, conditional on *Y*. The theoretical magnitude of the maximum additional bias has a more complicated form when *X* is partially observed because the imputation and analysis models are not the same. Again following the argument of Curnow *et al*. [16], the MI estimator will be unbiased only if unbiased estimates of all the imputation model parameters can be obtained using records with observed values of *X*. However, taking the imputation model parameter for *Y* as an example, and using a similar argument to the previous setting, we find that this parameter is biased, taking its maximum value of *β*_*XY*|*R*_ when the imputation model includes only *Y*, and *β*_*XY*|*R,Z*_ when the imputation model includes *Y* and *Z*, as the proportion of missing values tends to one.

In terms of the direct effect sizes and error variances,

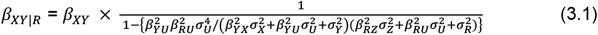

and

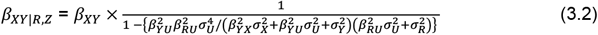

where the direct effect sizes are denoted by *β*_.._, *e.g. β*_*RU*_ denotes the direct effect of *U* on *R*, and the error variances are denoted by *σ*^2^, *e.g*.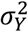 denotes the error variance of *Y*. Since |*β*_*XY*|*R,Z*_| > |*β*_*XY*|*R*_| > |*β*_*XY*_|, bias of the *Y* coefficient will be amplified when *Z* is also included as a predictor in the imputation model for *X* (see Supplementary Material, Section S3, for derivation of these results).

Due to its complexity in the setting in which *X* is partially observed, an expression for the theoretical magnitude of the maximum additional bias of the MI estimator is not derived here. However, we illustrate the effect on the MI estimate from including auxiliary variable *Z* in the imputation model by simulation (see Supplementary Material Section S4 for further details). Note that we refer to the MI or CRA “estimate” when describing simulation study results, rather than “estimator” (which we have used when describing algebraic results).

Figure 4 illustrates the impact of the direct effect sizes on the additional bias of the MI estimate when the imputation model includes *Z* as a predictor and *X* is partially observed (with 50% missing values). As before, the additional bias is plotted against *β*_*YX*_ and *β*_*RZ*_. The distribution of the maximum bias for each value of *β*_*YX*_ and *β*_*RZ*_ (represented as a box-plot) is due to the variation in *β*_*YU*_ and *β*_*RU*_. Figure 4 shows that the additional bias is small, regardless of the direct effect sizes. Results are similar if *X* is binary (see Supplementary Material, Figure S4).

**Figure 4.**
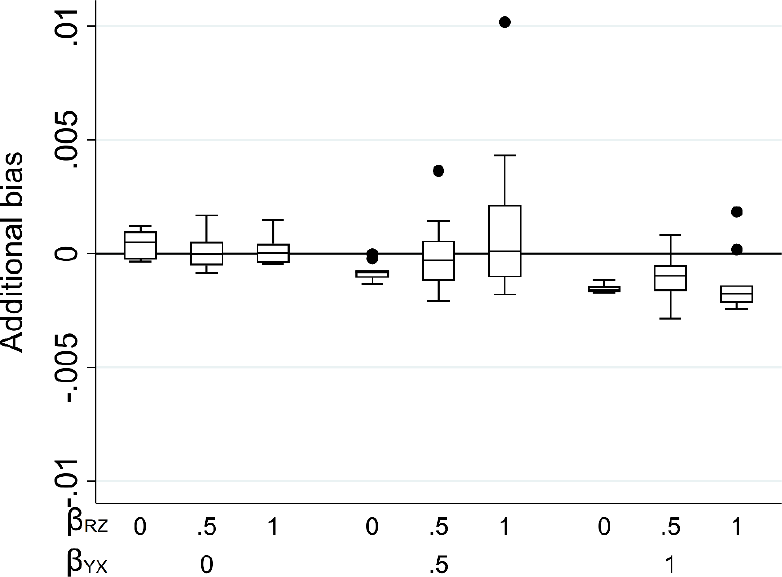
Additional bias of the MI estimate of β_YX_ when continuous exposure X is missing not at random, conditional on outcome Y, and the imputation model includes Y and an auxiliary variable Z that predicts missingness but not the missing values, with missingness related to Y via an unmeasured variable U. Results shown when 50% of values are missing, varying the direct effect sizes β_YX_, β_YU_, β_RU_, and β_RZ_. The distribution of additional bias in each box-plot is averaged over the values of β_YU_ and β_RU_.

## 4. Scenario 3. Additional bias of the MI estimator from including a predictor only of missingness in the imputation model when continuous outcome *Y* or continuous exposure *X* are partially observed and missingness is caused by both *X* and *Y*

Finally, we consider the setting in which the CRA and MI estimators are biased if either *Y* or *X* are partially observed: when *Y* and *X* directly cause missingness, as per Figure 5.

**Figure 5.**
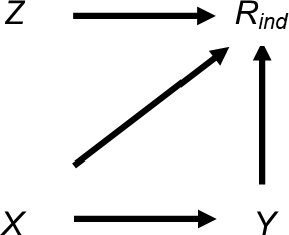
Directed acyclic graph depicting the relationship between outcome Y, exposure X, missingness indicator R_ind_, and potential auxiliary variable Z. Lines indicate related variables, with arrows indicating the direction of the relationship; absent lines represent variables with no direct causal relation

In this setting (given the same assumptions and using the same analysis model and MI method as in the previous scenarios), we can express both the maximum bias due to *Y* being MNAR (when using *X* as the predictor in the imputation model for *Y*) and the maximum additional bias (from including *Z* as well as *X* in the imputation model) in terms of the direct effect sizes and error variances (see Supplementary Material, Section S5, for derivation), as follows:

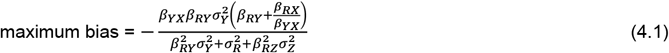

and

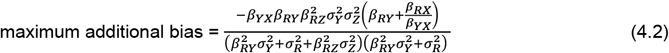

where the direct effect sizes are denoted by *β*.., *e.g. β*_*RY*_ denotes the direct effect of *Y* on *R*, and the error variances are denoted by *σ*^2^, *e.g*. 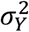 denotes the error variance of *Y*, as before. Note that in this setting, the maximum bias may be towards or away from zero, depending on the sign and magnitude of the direct effects, relative to the magnitude of the error variances. However, the maximum additional bias will always be in the same direction as the maximum bias, and will amplify the bias by a factor of

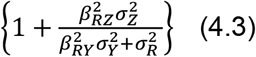

Note that this is identical to the amplification factor in Scenario 1 (although the bias due *Y* being MNAR in this setting may be greater or smaller than in Scenario 1, depending on the sign and magnitude of 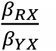).

When *X* is partially observed, the maximum additional bias of the *Y* coefficient in the imputation model for *X* (*i.e*. in addition to the bias due to *X* being MNAR) from including *Z* as a predictor in the imputation model is equal to:

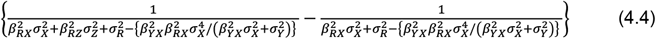

As in the previous scenario, we explored the effect of this additional bias on the MI estimate by simulation, due to the complexity of the theoretical expression for the maximum additional bias when *X* is partially observed (see Supplementary Material Section S4 for further details).

Figures 6 and 7 illustrate the impact of the direct effect sizes on the maximum additional bias when *Y* is partially observed and the additional bias when 50% of values of *X* are missing, respectively, when the imputation model includes *Z* as a predictor. The distribution of each box-plot is due to the variation in *β*_*RY*_ and *β*_*RX*_. When *Y* is partially observed, Figure 6 shows that the maximum additional bias is negative and increases in magnitude with the direct effect sizes (and is larger than in Scenario 1 for the same direct effect sizes). When *X* is partially observed, Figure 7 shows that the additional bias is negative and increases in magnitude with *β*_*RZ*_, as well as with *β*_*YX*_ when *β*_*YX*_ ≤ 0.5. However, the additional bias is smaller in magnitude when *β*_*YX*_ = 1. Results for partially observed *Y* are similar if *Y* is binary (see Supplementary Material, Figure S5). If *X* is binary, additional bias when *X* is partially observed increases with both *β*_*YX*_ and *β*_*RZ*_ (see Supplementary Material, Figure S6). The difference between results when *X* is continuous or binary may be due to the choice of distribution of *X* in each case: in the continuous case, *X* is normally distributed, with mean equal to 0 and variance equal to 1; in the binary case, *X* takes values of 0 or 1 with probability 0.5 (equivalent to a mean of 0.5 and a variance of 0.25).

**Figure 6.**
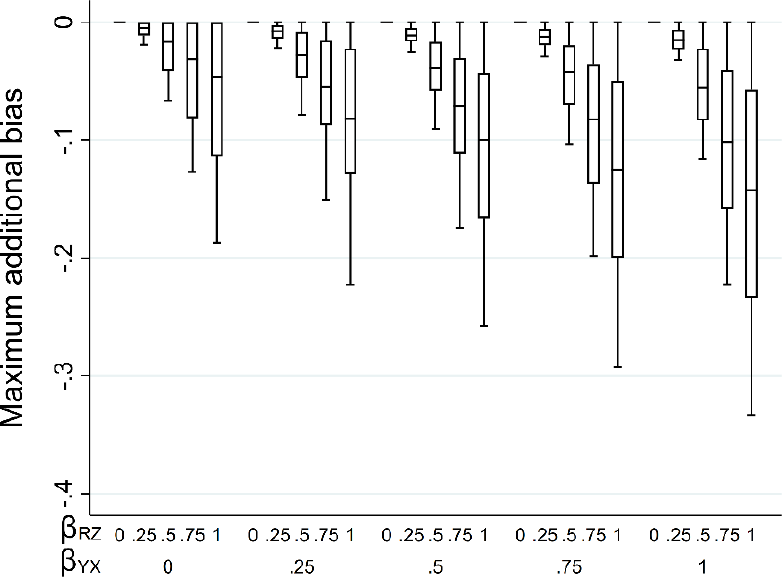
Maximum additional bias of the MI estimator of β_YX_ when continuous outcome Y is missing not at random, with missingness caused by Y and X, and the imputation model includes exposure X and a predictor of missingness but not the missing values, Z, varying the direct effect sizes β_YX_, β_RY_, β_RX_, and β_RZ_. The distribution of additional bias in each box-plot is averaged over the values of β_RY_ and β_RX_.

**Figure 7.**
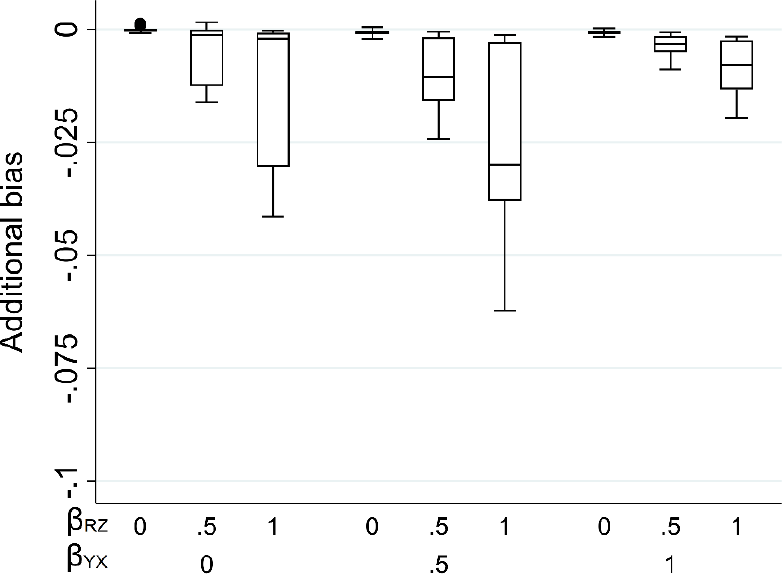
Additional bias of the MI estimate of β_YX_ when continuous exposure X is missing not at random, with missingness caused by Y and X, and the imputation model includes exposure Y and a predictor of missingness but not the missing values, Z. Results shown when 50% of values are missing, varying the direct effect sizes β_YX_, β_RY_, β_RX_, and β_RZ_. The distribution of additional bias in each box-plot is averaged over the values of β_RY_ and β_RX_.

## 5. Real data example

### 5.1. Methods

We illustrate this situation using data from the Avon Longitudinal Study of Parents and Children (ALSPAC). ALSPAC is a prospective study which recruited pregnant women with expected dates of delivery between 1^st^ April 1991 and 31^st^ December 1992, in the Bristol area of the UK [18, 19]. We use data from the initial recruitment phase, in which 14,541 pregnant women enrolled, resulting in 14,062 live births (13,988 alive at one year). This study uses data from all singletons and twins, where neither the mother nor child had withdrawn consent at the time of analysis (N=13,923). Children and their mothers have been followed up since birth through questionnaires, clinics, and linkage to routine datasets. ALSPAC has a searchable data dictionary: http://www.bristol.ac.uk/alspac/researchers/our-data/, describing all available data. Ethical approval was obtained from the ALSPAC Ethics and Law Committee and local research ethics committees. Informed consent for the use of data collected via questionnaires and clinics was obtained from participants following the recommendations of the ALSPAC Ethics and Law Committee at the time.

Here, our substantive model of interest is a linear regression of child’s IQ at age 15 years (*IQ15*) on breastfeeding duration (*bf*: categorised as never/< 3 months versus 3 months plus). Guided by previous studies [7, 12], we adjust for six confounders of the breastfeeding-IQ relationship, namely child’s sex, mother’s educational level (whether the child’s mother held a post-16 years qualification or not), mother’s occupational social class (professional, managerial, or non-manual skilled occupation vs. manual skilled, semi-skilled, or unskilled occupation), mother’s age and parity (number of previous births), and housing tenure (whether the family home was owned/mortgaged, privately rented, or rented from the local council or a housing association).

*IQ15* was not reported for 8913 (64%) participants in the study. Previous studies [7, 12] used linked educational attainment data to explore the missingness mechanism for *IQ15*. They found that *IQ15* was more likely to be missing for individuals with lower educational attainment (highly correlated with *IQ15*), which suggests *IQ15* is MNAR. We explore the consequences of performing MI when *IQ15* is likely to be MNAR, focusing particularly on the effect of including an auxiliary variable that is predictive of missingness but not the missing values of *IQ15*. Our chosen auxiliary variable is whether the mother smoked during the first trimester of pregnancy (*matsmok*). Note that there were also missing values for *bf*, confounders, and *matsmok*: *bf* was missing for 1406 (10%) individuals, values of one or more confounders were missing for 4394 (32%) individuals (although child’s sex and maternal age were fully observed), and *matsmok* was missing for 817 (6%) individuals. For simplicity, and purely for illustrative purposes, we assume that *bf*, confounders, and *matsmok* are MAR, conditional on the observed data.

Figure 8 depicts our hypothesised relationships between *IQ15, bf*, confounders (with confounders collectively denoted by *C* – for simplicity, we do not depict the relationships between individual confounders and/or missingness indicators for variables other than *IQ15*), potential auxiliary variable *matsmok*, and missingness indicator *R*_*IQ15*_ (a binary variable indicating whether *IQ15* is observed), plus related, unmeasured variable(s), *U* (*e.g*. markers of child’s behaviour). Here, we assume the setting is similar to that depicted in our theoretical Scenario 1 *i.e*. we assume missingness is caused by *IQ15* but not by our exposure, *bf*, or confounders (although we cannot rule out the possibility that missingness is caused by unmeasured variable(s) that are also related to *IQ15*). In Figure 8, black lines depict the relationships assumed in theoretical Scenario 1; grey lines depict additional relationships that are plausible in this real data setting.

**Figure 8.**
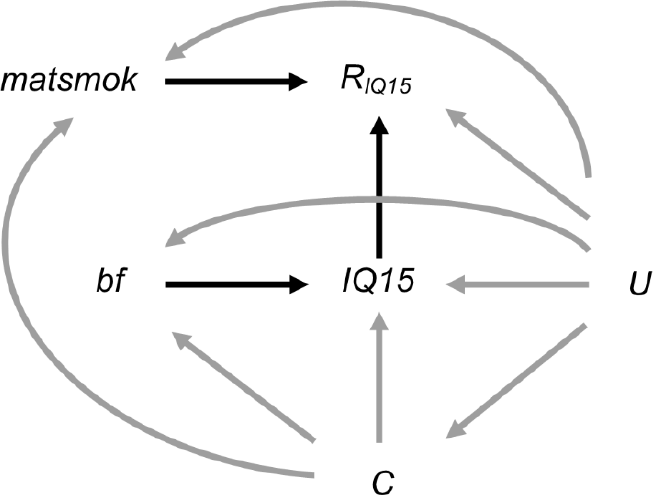
Directed acyclic graph depicting the relationship between child’s IQ at age 15 years (IQ15), duration of breastfeeding (bf), confounders of the IQ15-bf relationship (C), whether the mother smoked during the first trimester of pregnancy (matsmok), missingness indicator R_IQ15_ (a binary variable indicating whether IQ15 is observed), and unmeasured variable(s) U. Lines indicate related variables, with arrows indicating the direction of the relationship. Black lines depict the relationships assumed in theoretical Scenario 1; grey lines depict additional relationships that are plausible in our real data example; absent lines represent variables with no direct causal relationship.

We first assessed whether the hypothesised relationships between *IQ15, R*_*IQ15*_, *bf*, and *matsmok* were plausible by exploring the relationships in the observed data. We then applied our equation (Equation 2.4) for maximum bias amplification due to including predictor of missingness *matsmok* in the imputation model for *IQ15*. We assumed (without loss of generality) that *R* had a mean of zero and a variance of one. Therefore, we used the following version of Equation 2.4: maximum bias amplification = 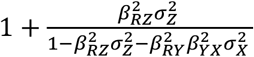 where, in our setting, *X* denotes *bf* and *Z* denotes *matsmok*. Parameter *β*_*RZ*_ and the product *β*_*RY*_*β*_*YX*_ were estimated as 0.6 × the coefficient for *matsmok* and *bf*, respectively, from a logistic regression of *R*_*IQ15*_ on *matsmok, bf*, and confounders (as before, multiplying by 0.6 to transform the parameters to the equivalent parameters from a probit regression of the underlying normal variable *R* [17]). We estimated 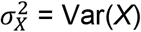 and 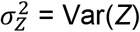 using the normal approximation to the binomial because *X* and *Z* were binary. We assumed that the estimates used in our maximum bias amplification equation were unbiased (which may not have been the case if there were unmeasured confounders of the relationship between *matsmok, bf*, and *R*_*IQ15*_).

We compared our estimate of the maximum bias amplification to both the CRA estimate and MI estimates using no auxiliary variables or using *matsmok* as an auxiliary variable. We used MI by chained equations [20] to impute missing values of *IQ15, bf*, confounders, and (where used) *matsmok*, including all other variables as predictors in the imputation model for each partially observed variable. We used a linear regression model to impute *IQ15*, logistic regression to impute *bf*, binary confounders, and *matsmok*, ordered logistic regression to impute parity, and multinomial logistic regression to impute housing tenure. We used 20 iterations in the imputation step and a large number of imputations (100) to ensure we obtained stable estimates of the exposure coefficient and its SE.

### 5.2. Results

The estimated association between *matsmok* and *IQ15*, adjusted for *bf* and confounders, was -0.79 (95% CI: -1.88, 0.31). The wide CI suggests that *matsmok* is only weakly predictive of *IQ15*, conditional on *bf* and confounders. We would expect some association between *matsmok* and *IQ15* in the observed data via the *matsmok* - *R*_*IQ15*_ - *IQ15* pathway *i.e*. due to collider/selection bias because we are conditioning on *R*_*IQ15*_. Estimates of the coefficient for *matsmok* and *bf* from a logistic regression of *R*_*IQ15*_ on *matsmok, bf*, and confounders, were -0.39 (95% CI: -0.51, -0.27) and 0.44 (95% CI: 0.35, 0.53), respectively which suggests that, conditional on the confounders, *matsmok* and *bf are* strongly predictive of missingness of *IQ15*. These results, combined with our prior knowledge of the data, suggest that inclusion of *matsmok* in the imputation model for *IQ15* may amplify any bias due to *IQ15* being MNAR.

Substituting values based on the observed data into our equation, we estimated that including *matsmok* in the imputation model for *IQ15* would amplify any bias in the *bf* coefficient due to *IQ15* being MNAR by 1% (towards the null). This result suggests that the magnitude of bias amplification is small in this particular setting.

Analysis results (Table 2) confirmed that MI estimates of the *bf* coefficient were very similar, regardless of whether auxiliary variable *matsmok* was used in the MI procedure, with the MI estimate based on *matsmok* smaller than the MI estimate based only on analysis model variables, as predicted by our equation. Both MI estimates were smaller than the CRA estimate. Based on the conclusions from previous studies [7, 12], both MI and CRA estimates under-estimate the true magnitude of the association. Using *matsmok*, a predictor of missingness but not of *IQ15* itself, as an auxiliary variable amplifies any bias, albeit the size of the bias amplification is small in this particular setting. Note that the directions of bias and bias amplification are consistent with results in the theoretical Scenario 1.

**Table 2.**
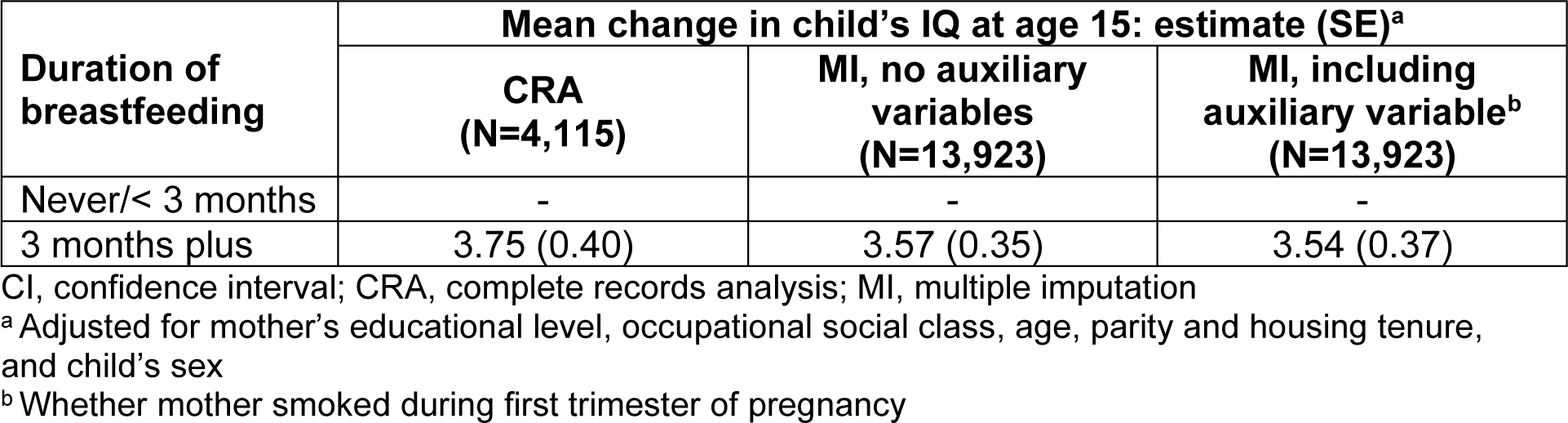
Relationship between child’s IQ at age 15 years and duration of breastfeeding, estimated using different analysis strategies.

| Duration of breastfeeding | Mean change in child's IQ at age 15: estimate (SE)<sup>a</sup> |  |  |
| --- | --- | --- | --- |
|  | CRA (N=4,115) | MI, no auxiliary variables (N=13,923) | MI, including auxiliary variable<sup>b</sup> (N=13,923) |
| Never/< 3 months | - | - | - |
| 3 months plus | 3.75 (0.40) | 3.57 (0.35) | 3.54 (0.37) |

CI, confidence interval; CRA, complete records analysis; MI, multiple imputation
<sup>a</sup> Adjusted for mother's educational level, occupational social class, age, parity and housing tenure, and child's sex
<sup>b</sup> Whether mother smoked during first trimester of pregnancy

## Discussion

In this paper, we quantify, algebraically and by simulation, the magnitude of the additional bias of the MI estimator, in addition to any bias due to data MNAR, from including a predictor of missingness but not the missing values themselves in the imputation model. We have derived algebraic expressions for the maximum additional bias when a continuous outcome is partially observed. We have demonstrated that if missingness is caused by the outcome, the additional bias can be substantial, relative to the magnitude of the exposure coefficient (and also if the outcome is binary). Furthermore, if missingness is caused by the outcome and the exposure, the additional bias can be even larger, when either the (continuous or binary) outcome or exposure is partially observed.

In addition, when a continuous analysis model outcome *Y* is partially observed and linear regression models are fitted (for both analysis and imputation), we have demonstrated algebraically the, perhaps surprising, result that if missingness is only related to *Y* via another variable *U* (where *U* causes *Y* and its missingness but is only related to exposure *X* and confounders via *Y*), then both CRA and MI will be unbiased even if *U* is not included in the analysis and imputation models. Furthermore, in this scenario, the bias of the MI estimate is likely to be small when binary *Y* (fitting a logistic regression model) or (continuous or binary) *X* is partially observed.

A strength of our approach is that we have considered a range of commonly-occurring scenarios, in which the partially observed variable is either the analysis model outcome or the exposure, as well as either continuous or binary. By using both algebra and simulation, we have been able to provide a detailed illustration of the magnitude of bias due to including auxiliary variables that only predict missingness, and how this is related to the magnitude and sign of individual assocations between exposure, outcome, auxiliary variables, and missingness. A limitation of our study is that we have only considered simple models, without interactions or non-linear relationships. However, since our general argument is based on a “missingness” DAG [21, 22], which does not make any distributional assumptions, our findings can be applied to more complex models (*e.g*. including an exposure-confounder interaction), to avoid using MI in a way which may increase bias (noting that the magnitude and direction of additional bias may be different from those suggested by our formulae in this case, particularly if either the analysis or missingness model includes interactions). A further limitation of our study is that in each of our scenarios, only a single variable has missing values. If multiple missingness is handled using MI by chained equations (as we did in the real data example), each imputation model only considers one variable to have missing values, as here. In this case, auxiliary variables should be considered separately for each imputation model, because an auxiliary variable may be predictive of one partially observed variable (and/or its missingness), but not another.

In summary, we conclude that, whilst auxiliary variables have the potential to improve precision of MI estimates and reduce bias due to data MNAR, the naïve and commonly used strategy of including all available auxiliary variables should be avoided. Any auxiliary variables that predict missingness but are only weakly predictive of the partially observed variable may cause additional bias, over and above any bias due to data MNAR. Although it is important to identify predictors of missingness to inform analysis strategy (*e.g*. to determine whether CRA is likely to be valid), our results show that such variables should not necessarily be included as predictors in the imputation models *i.e*. unless they also predict the partially observed variable. Given a choice of potential auxiliary variables, we recommend including the variables most predictive of the partially observed variable as auxiliary variables in the imputation model (in addition to all variables required for the analysis model) in order to minimise bias due to data MNAR. These variables can be identified through a combination of data exploration and consideration of the plausible casual diagrams and missingness mechanisms.

## Data Availability

Stata code to verify algebraic results, and also to generate and analyse the data as per the simulation studies is included in Supplementary Material, Section S6. Stata code to perform the real data analysis is included in Supplementary Material, Section S7. The real data are not publicly available due to privacy restrictions. Requests to access these datasets should be directed to.

## Acknowledgements

We are extremely grateful to all the families who took part in the ALSPAC study, the midwives for their help in recruiting them, and the whole ALSPAC team, which includes interviewers, computer and laboratory technicians, clerical workers, research scientists, volunteers, managers, receptionists and nurses.

