## Supplementary Material for "Multiple imputation assuming missing at random: auxiliary imputation variables that only predict missingness can increase bias due to data missing not at random"

*Section S1. Scenario 1. Derivation of the theoretical expression for the maximum additional bias of the MI estimator when a continuous outcome Y is missing not at random, with missingness caused by Y itself, when the imputation model includes an auxiliary variable Z that predicts missingness but not the missing values*

As per the main text, we assume that $Y$, $X$, $Z$, and $R$ are normally distributed, such that $Y = \beta_{YX}X + \varepsilon_Y$, where $\varepsilon_Y \sim N(0, \sigma_Y^2)$, $X \sim N(\mu_X, \sigma_X^2)$, $Z \sim N(\mu_Z, \sigma_Z^2)$, and $R = \beta_{RY}Y + \beta_{RZ}Z + \varepsilon_R$ where $\varepsilon_R \sim N(0, \sigma_R^2)$. We further assume that each of $Y$ and $R$ is a linear combination of the variables causing it plus an error term (with $X$ and $Z$ having no direct causes), with no interactions, all errors uncorrelated, no model mis-specification, and no measurement error, and that an ordinary least squares (OLS) estimator is used to obtain estimates in both analysis and imputation models.

Then, using standard results, the joint distribution of $Y$, $X$, $Z$, and $R$, $f(Y, X, Z, R)$, is multivariate normal with covariance matrix $\Sigma$, where:

$$\Sigma = \begin{pmatrix} \beta_{YX}^2\sigma_X^2 + \sigma_Y^2 & \beta_{YX}\sigma_X^2 & 0 & \beta_{RY}(\beta_{YX}^2\sigma_X^2 + \sigma_Y^2) \\ \beta_{YX}\sigma_X^2 & \sigma_X^2 & 0 & \beta_{YX}\beta_{RY}\sigma_X^2 \\ 0 & 0 & \sigma_Z^2 & \beta_{RZ}\sigma_Z^2 \\ \beta_{RY}(\beta_{YX}^2\sigma_X^2 + \sigma_Y^2) & \beta_{YX}\beta_{RY}\sigma_X^2 & \beta_{RZ}\sigma_Z^2 & Var(R) \end{pmatrix}$$

where $Var(R) = \beta_{RZ}^2\sigma_Z^2 + \beta_{RY}^2(\beta_{YX}^2\sigma_X^2 + \sigma_Y^2) + \sigma_R^2$

Then, again using standard results, the joint conditional distribution of *Y*, *X*, and *Z,* given *R*, *f*(*Y*, *X*, *Z* | *R*=*r*), is multivariate normal with covariance matrix ∑*, where:

$$\Sigma^* = \begin{pmatrix} \beta_{YX}^2\sigma_X^2 + \sigma_Y^2 & \beta_{YX}\sigma_X^2 & 0 \\ \beta_{YX}\sigma_X^2 & \sigma_X^2 & 0 \\ 0 & 0 & \sigma_Z^2 \end{pmatrix} - \frac{1}{Var(R)} \times$$
$$\begin{pmatrix} \beta_{RY}(\beta_{YX}^2\sigma_X^2 + \sigma_Y^2) \\ \beta_{YX}\beta_{RY}\sigma_X^2 \\ \beta_{RZ}\sigma_Z^2 \end{pmatrix} \begin{pmatrix} \beta_{RY}(\beta_{YX}^2\sigma_X^2 + \sigma_Y^2) & \beta_{YX}\beta_{RY}\sigma_X^2 & \beta_{RZ}\sigma_Z^2 \end{pmatrix}$$

$$= \begin{pmatrix} \beta_{YX}^2\sigma_X^2 + \sigma_Y^2 & \beta_{YX}\sigma_X^2 & 0 \\ \beta_{YX}\sigma_X^2 & \sigma_X^2 & 0 \\ 0 & 0 & \sigma_Z^2 \end{pmatrix} - \frac{1}{Var(R)} \times$$
$$\begin{pmatrix} \beta_{RY}^2(\beta_{YX}^2\sigma_X^2 + \sigma_Y^2)^2 & \beta_{YX}\beta_{RY}^2(\beta_{YX}^2\sigma_X^2 + \sigma_Y^2)\sigma_X^2 & \beta_{RY}\beta_{RZ}(\beta_{YX}^2\sigma_X^2 + \sigma_Y^2)\sigma_Z^2 \\ \beta_{YX}\beta_{RY}^2(\beta_{YX}^2\sigma_X^2 + \sigma_Y^2)\sigma_X^2 & \beta_{YX}^2\beta_{RY}^2\sigma_X^4 & \beta_{YX}\beta_{RY}\beta_{RZ}\sigma_X^2\sigma_Z^2 \\ \beta_{RY}\beta_{RZ}(\beta_{YX}^2\sigma_X^2 + \sigma_Y^2)\sigma_Z^2 & \beta_{YX}\beta_{RY}\beta_{RZ}\sigma_X^2\sigma_Z^2 & \beta_{RZ}^2\sigma_Z^4 \end{pmatrix}$$

Hence, $\beta_{YX|R} = \frac{Cov(X,Y|R)}{Var(X|R)} = \frac{\beta_{YX}\sigma_X^2\{Var(R) - \beta_{RY}^2(\beta_{YX}^2\sigma_X^2+\sigma_Y^2)\}}{\sigma_X^2\{Var(R) - \beta_{YX}^2\beta_{RY}^2\sigma_X^2\}}$

$= \frac{\beta_{YX}\{\beta_{RZ}^2\sigma_Z^2+\beta_{YX}^2\beta_{RY}^2\sigma_X^2+\beta_{RY}^2\sigma_Y^2+\sigma_R^2 - \beta_{YX}^2\beta_{RY}^2\sigma_X^2 - \beta_{RY}^2\sigma_Y^2\}}{\beta_{RZ}^2\sigma_Z^2+\beta_{YX}^2\beta_{RY}^2\sigma_X^2+\beta_{RY}^2\sigma_Y^2+\sigma_R^2 - \beta_{YX}^2\beta_{RY}^2\sigma_X^2}$

$= \frac{\beta_{YX}\{\beta_{RZ}^2\sigma_Z^2 + \sigma_R^2\}}{\beta_{RZ}^2\sigma_Z^2+\beta_{RY}^2\sigma_Y^2+\sigma_R^2}$

$= \beta_{YX} \times \left\{1 - \frac{\beta_{RY}^2\sigma_Y^2}{\beta_{RY}^2\sigma_Y^2+\beta_{RZ}^2\sigma_Z^2+\sigma_R^2}\right\}$ as per Formula 2.1 in the main text.

Using a similar approach, the joint conditional distribution of *Y* and *X* given *Z* and *R*, *f*(*Y*, *X* | *Z*=z, *R*=r), is multivariate normal with covariance matrix ∑**, where

$$\Sigma^{**} = \begin{pmatrix} \beta_{YX}^2\sigma_X^2 + \sigma_Y^2 & \beta_{YX}\sigma_X^2 \\ \beta_{YX}\sigma_X^2 & \sigma_X^2 \end{pmatrix} - \frac{1}{\sigma_Z^2 Var(R) - \beta_{RZ}^2\sigma_Z^4} \times$$
$$\begin{pmatrix} 0 & \beta_{RY}(\beta_{YX}^2\sigma_X^2 + \sigma_Y^2) \\ 0 & \beta_{YX}\beta_{RY}\sigma_X^2 \end{pmatrix} \begin{pmatrix} Var(R) & -\beta_{RZ}\sigma_Z^2 \\ -\beta_{RZ}\sigma_Z^2 & \sigma_Z^2 \end{pmatrix} \begin{pmatrix} 0 & 0 \\ \beta_{RY}(\beta_{YX}^2\sigma_X^2 + \sigma_Y^2) & \beta_{YX}\beta_{RY}\sigma_X^2 \end{pmatrix}$$

$$= \begin{pmatrix} \beta_{YX}^2\sigma_X^2 + \sigma_Y^2 & \beta_{YX}\sigma_X^2 \\ \beta_{YX}\sigma_X^2 & \sigma_X^2 \end{pmatrix} - \frac{1}{\sigma_Z^2 Var(R) - \beta_{RZ}^2\sigma_Z^4} \times$$

$$\begin{pmatrix}0 & \beta_{RY}(\beta_{YX}^2\sigma_X^2+\sigma_Y^2)\\ 0 & \beta_{YX}\beta_{RY}\sigma_X^2\end{pmatrix}\begin{pmatrix}-\beta_{RZ}\beta_{RY}(\beta_{YX}^2\sigma_X^2+\sigma_Y^2)\sigma_Z^2 & -\beta_{YX}\beta_{RZ}\beta_{RY}\sigma_X^2\sigma_Z^2\\ \beta_{RY}(\beta_{YX}^2\sigma_X^2+\sigma_Y^2)\sigma_Z^2 & \beta_{YX}\beta_{RY}\sigma_X^2\sigma_Z^2\end{pmatrix}$$

$$= \begin{pmatrix}\beta_{YX}^2\sigma_X^2+\sigma_Y^2 & \beta_{YX}\sigma_X^2\\ \beta_{YX}\sigma_X^2 & \sigma_X^2\end{pmatrix} - \frac{1}{\sigma_Z^2 Var(R)-\beta_{RZ}^2\sigma_Z^4}\times$$

$$\begin{pmatrix}\beta_{RY}^2(\beta_{YX}^2\sigma_X^2+\sigma_Y^2)^2\sigma_Z^2 & \beta_{YX}\beta_{RY}^2(\beta_{YX}^2\sigma_X^2+\sigma_Y^2)\sigma_X^2\sigma_Z^2\\ \beta_{YX}\beta_{RY}^2(\beta_{YX}^2\sigma_X^2+\sigma_Y^2)\sigma_X^2\sigma_Z^2 & \beta_{YX}^2\beta_{RY}^2\sigma_X^4\sigma_Z^2\end{pmatrix}$$

Hence, $\beta_{YX|Z,R} = \frac{Cov(X,Y|Z,R)}{Var(X|Z,R)} = \frac{\beta_{YX}\sigma_X^2\sigma_Z^2\{Var(R)-\beta_{RZ}^2\sigma_Z^2-\beta_{RY}^2(\beta_{YX}^2\sigma_X^2+\sigma_Y^2)\}}{\sigma_X^2\sigma_Z^2\{Var(R)-\beta_{RZ}^2\sigma_Z^2-\beta_{YX}^2\beta_{RY}^2\sigma_X^2\}}$

$$= \beta_{YX}\times\frac{\beta_{RZ}^2\sigma_Z^2+\beta_{RY}^2(\beta_{YX}^2\sigma_X^2+\sigma_Y^2)+\sigma_R^2-\beta_{RZ}^2\sigma_Z^2-\beta_{RY}^2(\beta_{YX}^2\sigma_X^2+\sigma_Y^2)}{\beta_{RZ}^2\sigma_Z^2+\beta_{RY}^2(\beta_{YX}^2\sigma_X^2+\sigma_Y^2)+\sigma_R^2-\beta_{RZ}^2\sigma_Z^2-\beta_{YX}^2\beta_{RY}^2\sigma_X^2}$$

$= \beta_{YX}\times\frac{\sigma_R^2}{\beta_{RY}^2\sigma_Y^2+\sigma_R^2} = \beta_{YX}\times\left\{1-\frac{\beta_{RY}^2\sigma_Y^2}{\beta_{RY}^2\sigma_Y^2+\sigma_R^2}\right\}$ as per Formula 2.2 in the main text.

Recall that $\beta_{YX|R} = \beta_{YX}\times\left\{1-\frac{\beta_{RY}^2\sigma_Y^2}{\beta_{RY}^2\sigma_Y^2+\sigma_R^2+\beta_{RZ}^2\sigma_Z^2}\right\}$. Thus, the maximum bias of the MI estimator due to *Y* being MNAR (using only *X* as a predictor in the imputation model for *Y*) is $-\frac{\beta_{YX}\beta_{RY}^2\sigma_Y^2}{\beta_{RY}^2\sigma_Y^2+\sigma_R^2+\beta_{RZ}^2\sigma_Z^2}$. The maximum additional bias of the MI estimator (*i.e.* in addition to the bias due to *Y* being MNAR) from including *Z* as a predictor in the imputation model is $\beta_{YX}\beta_{RY}^2\sigma_Y^2\times\left\{\frac{1}{\beta_{RY}^2\sigma_Y^2+\sigma_R^2+\beta_{RZ}^2\sigma_Z^2}-\frac{1}{\beta_{RY}^2\sigma_Y^2+\sigma_R^2}\right\} = \frac{-\beta_{YX}\beta_{RY}^2\beta_{RZ}^2\sigma_Y^2\sigma_Z^2}{(\beta_{RY}^2\sigma_Y^2+\sigma_R^2+\beta_{RZ}^2\sigma_Z^2)(\beta_{RY}^2\sigma_Y^2+\sigma_R^2)}$ as per Formula 2.3 in the main text. Or in other words, if bias amplification is defined as the bias of $\beta_{YX|Z,R}$ divided by the bias of $\beta_{YX|R}$, then maximum bias amplification = $\frac{-\beta_{YX}\beta_{RY}^2\sigma_Y^2}{\beta_{RY}^2\sigma_Y^2+\sigma_R^2} / \frac{-\beta_{YX}\beta_{RY}^2\sigma_Y^2}{\beta_{RY}^2\sigma_Y^2+\sigma_R^2+\beta_{RZ}^2\sigma_Z^2} = \frac{\beta_{RY}^2\sigma_Y^2+\sigma_R^2+\beta_{RZ}^2\sigma_Z^2}{\beta_{RY}^2\sigma_Y^2+\sigma_R^2} = 1+\frac{\beta_{RZ}^2\sigma_Z^2}{\beta_{RY}^2\sigma_Y^2+\sigma_R^2}$, that is, the maximum bias due to *Y* being MNAR is amplified by a factor of $\left\{1+\frac{\beta_{RZ}^2\sigma_Z^2}{\beta_{RY}^2\sigma_Y^2+\sigma_R^2}\right\}$ when *Z* is included in the imputation model for *Y*, as per Formula 2.4 in the main text.

*Section S2. Scenario 1. Verification of the theoretical expression for the maximum additional bias of the MI estimator when a continuous outcome Y is missing not at random, with missingness caused by Y itself, when the imputation model includes an auxiliary variable Z that predicts missingness but not the missing values*

The theoretical expressions for the maximum bias of the MI estimator due to Y being MNAR (when the imputation model includes only *X*), as well as the maximum additional bias of the MI estimator when the imputation model includes *X* and *Z*, were verified using simulation. We used 1000 simulations, and each simulated dataset contained 100,000 observations. In each simulated dataset, the values of each coefficient ($\beta_{YX}$, $\beta_{RZ}$, *etc.*) and each error variance ($\sigma_X^2$, $\sigma_Z^2$, *etc.*) were sampled from a uniform distribution *U*(0, 2). For simplicity, $\mu_X$

and $\mu_Z$ were set equal to zero (note that the formulas do not depend on these parameters). Data were then generated using the specified models for *Y*, *X*, *Z*, and *R*, as per Section S1, above. The two bias quantities were calculated using the theoretical expressions (Formulas 2.1 and 2.3 in the main text – note that verification of these two formulas also implicitly verifies Formulas 2.2 and 2.4). They were also estimated empirically by calculating the difference in the *X* coefficient when fitting a linear regression of (i) *Y* on *X*, (ii) *Y* on *X*, conditional on *R*, and (iii) *Y* on *X*, conditional on *R* and *Z* (with the difference between the coefficient from models (i) and (ii) used to estimate the maximum bias, and the difference between the coefficient from models (ii) and (iii) used to estimate the maximum additional bias).

The median difference between the theoretical and empirical values of maximum bias and maximum additional bias was 0.000 ($5^{th}$ – $95^{th}$ percentile: -0.007 - 0.009) and 0.000 ($5^{th}$ – $95^{th}$ percentile: -0.004 - 0.005), respectively. Therefore, we conclude that the theoretical expressions are correct.

*Section S3. Scenario 2. Derivation of the theoretical expression for the maximum additional bias when a continuous outcome Y or a continuous exposure X is partially observed, with missingness related to Y via an unmeasured variable U, when the imputation model includes an auxiliary variable Z that predicts missingness but not the missing values*

As per Figure 3 in the main text, we assume that *Y*, *X*, *Z*, *U*, and *R* are normally distributed, such that $Y = \beta_{YX}X + \beta_{YU}U + \varepsilon_Y$, where $\varepsilon_Y$ ~ N(0, $\sigma_Y^2$), *X* ~ N($\mu_X$, $\sigma_X^2$), *Z* ~ N($\mu_Z$, $\sigma_Z^2$), *U* ~ N($\mu_U$, $\sigma_U^2$), and $R = \beta_{RU}U + \beta_{RZ}Z + \varepsilon_R$, where $\varepsilon_R$ ~ N(0, $\sigma_R^2$). We once again assume that each of *Y* and *R* is a linear combination of the variables causing it plus an error term (with *X*, *Z*, and *U* having no direct causes), with no interactions, all errors uncorrelated, no model mis-specification, and no measurement error, and that an ordinary least squares (OLS) estimator is used to obtain estimates in both analysis and imputation models.

Then, using standard results, the joint distribution of *Y*, *X*, *Z*, *U*, and *R*, *f*(*Y*, *X*, *Z*, U, R), is multivariate normal with covariance matrix ∑, where:

$$\sum = \begin{pmatrix} \beta_{YX}^2\sigma_X^2 + \beta_{YU}^2\sigma_U^2 + \sigma_Y^2 & \beta_{YX}\sigma_X^2 & 0 & \beta_{YU}\sigma_U^2 & \beta_{YU}\beta_{RU}\sigma_U^2 \\ \beta_{YX}\sigma_X^2 & \sigma_X^2 & 0 & 0 & 0 \\ 0 & 0 & \sigma_Z^2 & 0 & \beta_{RZ}\sigma_Z^2 \\ \beta_{YU}\sigma_U^2 & 0 & 0 & \sigma_U^2 & \beta_{RU}\sigma_U^2 \\ \beta_{YU}\beta_{RU}\sigma_U^2 & 0 & \beta_{RZ}\sigma_Z^2 & \beta_{RU}\sigma_U^2 & Var(R) \end{pmatrix}$$

where $Var(R) = \beta_{RZ}^2\sigma_Z^2 + \beta_{RU}^2\sigma_U^2 + \sigma_R^2$

Then, again using standard results, the joint conditional distribution of *Y*, *X*, *Z*, and *U* given *R*, *f*(*Y*, *X*, *Z*, *U* | *R*=r), is multivariate normal with covariance matrix ∑*, where:

$$\Sigma^* = \begin{pmatrix} \beta_{YX}^2\sigma_X^2 + \beta_{YU}^2\sigma_U^2 + \sigma_Y^2 & \beta_{YX}\sigma_X^2 & 0 & \beta_{YU}\sigma_U^2 \\ \beta_{YX}\sigma_X^2 & \sigma_X^2 & 0 & 0 \\ 0 & 0 & \sigma_Z^2 & 0 \\ \beta_{YU}\sigma_U^2 & 0 & 0 & \sigma_U^2 \end{pmatrix} - \frac{1}{Var(R)} \times \begin{pmatrix} \beta_{YU}\beta_{RU}\sigma_U^2 \\ 0 \\ \beta_{RZ}\sigma_Z^2 \\ \beta_{RU}\sigma_U^2 \end{pmatrix} (\beta_{YU}\beta_{RU}\sigma_U^2 \quad 0 \quad \beta_{RZ}\sigma_Z^2 \quad \beta_{RU}\sigma_U^2)$$

$$= \begin{pmatrix} \beta_{YX}^2\sigma_X^2 + \beta_{YU}^2\sigma_U^2 + \sigma_Y^2 & \beta_{YX}\sigma_X^2 & 0 & \beta_{YU}\sigma_U^2 \\ \beta_{YX}\sigma_X^2 & \sigma_X^2 & 0 & 0 \\ 0 & 0 & \sigma_Z^2 & 0 \\ \beta_{YU}\sigma_U^2 & 0 & 0 & \sigma_U^2 \end{pmatrix} - \frac{1}{Var(R)} \times \begin{pmatrix} \beta_{YU}^2\beta_{RU}^2\sigma_U^4 & 0 & \beta_{YU}\beta_{RU}\beta_{RZ}\sigma_U^2\sigma_Z^2 & \beta_{YU}\beta_{RU}^2\sigma_U^4 \\ 0 & 0 & 0 & 0 \\ \beta_{YU}\beta_{RU}\beta_{RZ}\sigma_U^2\sigma_Z^2 & 0 & \beta_{RZ}^2\sigma_Z^4 & \beta_{RU}\beta_{RZ}\sigma_U^2\sigma_Z^2 \\ \beta_{YU}\beta_{RU}^2\sigma_U^4 & 0 & \beta_{RU}\beta_{RZ}\sigma_U^2\sigma_Z^2 & \beta_{RU}^2\sigma_U^4 \end{pmatrix}$$

Hence, $\beta_{YX|R} = \frac{Cov(X,Y|R)}{Var(X|R)} = \frac{\beta_{YX}\sigma_X^2}{\sigma_X^2} = \beta_{YX}$, which means that both CRA and MI estimators are unbiased when *Y* is partially observed.

However, note that $\beta_{XY|R} = \frac{Cov(X,Y|R)}{Var(Y|R)}$

$$= \frac{\beta_{YX}\sigma_X^2}{\beta_{YX}^2\sigma_X^2 + \beta_{YU}^2\sigma_U^2 + \sigma_Y^2 - \beta_{YU}^2\beta_{RU}^2\sigma_U^4/Var(R)}$$

$$= \frac{\beta_{YX}\sigma_X^2}{\beta_{YX}^2\sigma_X^2 + \beta_{YU}^2\sigma_U^2 + \sigma_Y^2} \times \frac{1}{1 - \beta_{YU}^2\beta_{RU}^2\sigma_U^4/\{(\beta_{YX}^2\sigma_X^2 + \beta_{YU}^2\sigma_U^2 + \sigma_Y^2)Var(R)\}}$$

$$= \beta_{XY} \times \frac{1}{1 - \{\beta_{YU}^2\beta_{RU}^2\sigma_U^4/(\beta_{YX}^2\sigma_X^2 + \beta_{YU}^2\sigma_U^2 + \sigma_Y^2)(\beta_{RZ}^2\sigma_Z^2 + \beta_{RU}^2\sigma_U^2 + \sigma_R^2)\}}$$

as per Formula 3.1 in the main text.

Using a similar approach, the joint conditional distribution of *Y*, *X*, and *U* given *Z* and *R*, *f*(*Y*, *X*, *U* | *Z*=z, *R*=r), is multivariate normal with covariance matrix ∑**, where

$$\Sigma^{**} = \begin{pmatrix} \beta_{YX}^2\sigma_X^2 + \beta_{YU}^2\sigma_U^2 + \sigma_Y^2 & \beta_{YX}\sigma_X^2 & \beta_{YU}\sigma_U^2 \\ \beta_{YX}\sigma_X^2 & \sigma_X^2 & 0 \\ \beta_{YU}\sigma_U^2 & 0 & \sigma_U^2 \end{pmatrix} - \frac{1}{\sigma_Z^2 Var(R) - \beta_{RZ}^2\sigma_Z^4} \times$$

$$\begin{pmatrix} 0 & \beta_{YU}\beta_{RU}\sigma_U^2 \\ 0 & 0 \\ 0 & \beta_{RU}\sigma_U^2 \end{pmatrix} \begin{pmatrix} Var(R) & -\beta_{RZ}\sigma_Z^2 \\ -\beta_{RZ}\sigma_Z^2 & \sigma_Z^2 \end{pmatrix} \begin{pmatrix} 0 & 0 & 0 \\ \beta_{YU}\beta_{RU}\sigma_U^2 & 0 & \beta_{RU}\sigma_U^2 \end{pmatrix}$$

$$= \begin{pmatrix} \beta_{YX}^2\sigma_X^2 + \beta_{YU}^2\sigma_U^2 + \sigma_Y^2 & \beta_{YX}\sigma_X^2 & \beta_{YU}\sigma_U^2 \\ \beta_{YX}\sigma_X^2 & \sigma_X^2 & 0 \\ \beta_{YU}\sigma_U^2 & 0 & \sigma_U^2 \end{pmatrix} - \frac{1}{\sigma_Z^2 Var(R) - \beta_{RZ}^2\sigma_Z^4} \times$$

$$\begin{pmatrix} 0 & \beta_{YU}\beta_{RU}\sigma_U^2 \\ 0 & 0 \\ 0 & \beta_{RU}\sigma_U^2 \end{pmatrix} \begin{pmatrix} -\beta_{RZ}\beta_{YU}\beta_{RU}\sigma_Z^2\sigma_U^2 & 0 & -\beta_{RZ}\beta_{RU}\sigma_Z^2\sigma_U^2 \\ \beta_{YU}\beta_{RU}\sigma_Z^2\sigma_U^2 & 0 & \beta_{RU}\sigma_Z^2\sigma_U^2 \end{pmatrix}$$

$$= \begin{pmatrix} \beta_{YX}^2\sigma_X^2 + \beta_{YU}^2\sigma_U^2 + \sigma_Y^2 & \beta_{YX}\sigma_X^2 & \beta_{YU}\sigma_U^2 \\ \beta_{YX}\sigma_X^2 & \sigma_X^2 & 0 \\ \beta_{YU}\sigma_U^2 & 0 & \sigma_U^2 \end{pmatrix} - \frac{1}{\sigma_Z^2 Var(R) - \beta_{RZ}^2\sigma_Z^4} \times$$

$$\begin{pmatrix} \beta_{YU}^2\beta_{RU}^2\sigma_Z^2\sigma_U^4 & 0 & \beta_{YU}\beta_{RU}^2\sigma_Z^2\sigma_U^4 \\ 0 & 0 & 0 \\ \beta_{YU}\beta_{RU}^2\sigma_Z^2\sigma_U^4 & 0 & \beta_{RU}^2\sigma_Z^2\sigma_U^4 \end{pmatrix}$$

Hence, $\beta_{YX|R,Z} = \frac{Cov(X,Y|R,Z)}{Var(X|R,Z)} = \frac{\beta_{YX}\sigma_X^2}{\sigma_X^2} = \beta_{YX}$. Therefore, the MI estimator will be unbiased when *Y* is partially observed, regardless of whether *Z* is included in the imputation model for *Y*.

However, $\beta_{XY|R,Z} = \frac{Cov(X,Y|R,Z)}{Var(Y|R,Z)}$

$$= \frac{\beta_{YX}\sigma_X^2}{\beta_{YX}^2\sigma_X^2 + \beta_{YU}^2\sigma_U^2 + \sigma_Y^2 - \beta_{YU}^2\beta_{RU}^2\sigma_Z^2\sigma_U^4/(\sigma_Z^2 Var(R) - \beta_{RZ}^2\sigma_Z^4)}$$

$$= \frac{\beta_{YX}\sigma_X^2}{\beta_{YX}^2\sigma_X^2 + \beta_{YU}^2\sigma_U^2 + \sigma_Y^2}$$

$$\times \frac{1}{1 - \{\beta_{YU}^2\beta_{RU}^2\sigma_U^4/(\beta_{YX}^2\sigma_X^2 + \beta_{YU}^2\sigma_U^2 + \sigma_Y^2)(\beta_{RZ}^2\sigma_Z^2 + \beta_{RU}^2\sigma_U^2 + \sigma_R^2 - \beta_{RZ}^2\sigma_Z^2)\}}$$

$$= \beta_{XY} \times \frac{1}{1 - \{\beta_{YU}^2\beta_{RU}^2\sigma_U^4/(\beta_{YX}^2\sigma_X^2 + \beta_{YU}^2\sigma_U^2 + \sigma_Y^2)(\beta_{RU}^2\sigma_U^2 + \sigma_R^2)\}}$$

as per Formula 3.2 in the main text.

Recall that $\beta_{XY|R} = \beta_{XY} \times \frac{1}{1-\{\beta_{YU}^2\beta_{RU}^2\sigma_U^4/(\beta_{YX}^2\sigma_X^2+\beta_{YU}^2\sigma_U^2+\sigma_Y^2)(\beta_{RZ}^2\sigma_Z^2+\beta_{RU}^2\sigma_U^2+\sigma_R^2)\}} = \beta_{XY} \times \frac{1}{1-\alpha^2}$

where $\alpha^2 = \frac{\beta_{YU}^2\beta_{RU}^2\sigma_U^4}{(\beta_{YX}^2\sigma_X^2+\beta_{YU}^2\sigma_U^2+\sigma_Y^2)(\beta_{RZ}^2\sigma_Z^2+\beta_{RU}^2\sigma_U^2+\sigma_R^2)} = \frac{\beta_{YU}^2\beta_{RU}^2\sigma_U^4}{\beta_{YU}^2\beta_{RU}^2\sigma_U^4 + other\ positive\ terms}$

Since 0 < $\alpha^2$ < 1 (assuming Cov(*Y*,*R*) ≠ 0), $|\beta_{XY|R}|$ is always greater than $|\beta_{XY}|$.

Furthermore, $\beta_{XY|R,Z} = \beta_{XY} \times \frac{1}{1-\tau^2\alpha^2}$ where $\alpha^2$ is defined as above and $\tau^2 = \frac{\beta_{RZ}^2\sigma_Z^2+\beta_{RU}^2\sigma_U^2+\sigma_R^2}{\beta_{RU}^2\sigma_U^2+\sigma_R^2}$

Since $\tau^2$ > 1 (assuming Cov(*Z*,*R*) ≠ 0), $\tau^2\alpha^2 > \alpha^2 > 0$.

Also, $\tau^2\alpha^2 = \frac{\beta_{YU}^2\beta_{RU}^2\sigma_U^4}{(\beta_{YX}^2\sigma_X^2+\beta_{YU}^2\sigma_U^2+\sigma_Y^2)(\beta_{RZ}^2\sigma_Z^2+\beta_{RU}^2\sigma_U^2+\sigma_R^2)} \times \frac{\beta_{RZ}^2\sigma_Z^2+\beta_{RU}^2\sigma_U^2+\sigma_R^2}{\beta_{RU}^2\sigma_U^2+\sigma_R^2}$

$= \frac{\beta_{YU}^2\beta_{RU}^2\sigma_U^4}{(\beta_{YX}^2\sigma_X^2+\beta_{YU}^2\sigma_U^2+\sigma_Y^2)(\beta_{RU}^2\sigma_U^2+\sigma_R^2)} = \frac{\beta_{YU}^2\beta_{RU}^2\sigma_U^4}{\beta_{YU}^2\beta_{RU}^2\sigma_U^4 + other\ positive\ terms} < 1$

Therefore, 0 < $\alpha^2$ < $\tau^2\alpha^2$ < 1, and hence, $|\beta_{XY|R,Z}|$ > $|\beta_{XY|R}|$ > $|\beta_{XY}|$ and so bias of the *Y* coefficient will be amplified when *Z* is also included as a predictor in the imputation model for *X* (as well as *Y*) in the main text.

Using the same approach as in Section S2, the theoretical expression for $\beta_{XY|R}$ (*i.e.* the magnitude of the imputation model parameter $\alpha_1^{OBS}$ when the imputation model includes only *Y* and the proportion of missing values tends to one), as well as for $\beta_{XY|R,Z}$ (*i.e.* the magnitude of the imputation model parameter $\alpha_1^{OBS}$ when the imputation model includes *Y* and *Z* and the proportion of missing values tends to one), were verified using simulation. We also verified that $\beta_{YX|R} = \beta_{YX|R,Z} = \beta_{YX}$ when *Y* was partially observed (*i.e.* that there was no bias using either imputation model in this setting). We used 1000 simulations, and each simulated dataset contained 100,000 observations. In each simulated dataset, the values of each coefficient ($\beta_{YX}$, $\beta_{RZ}$, *etc.*) and each error variance ($\sigma_X^2$, $\sigma_Z^2$, *etc.*) were sampled from a uniform distribution *U*(0, 2). For simplicity, $\mu_X$, $\mu_Z$, and $\mu_U$ were set equal to zero (note that the formulas do not depend on these parameters). Data were then generated using the models for *Y*, *X*, *Z*, *U*, and *R*, specified above. $\beta_{XY|R}$ and $\beta_{XY|R,Z}$ were calculated using the theoretical expressions above. They were also estimated empirically by calculating the *Y* coefficient when fitting a linear regression of (i) *X* on *Y*, conditional on *R*, and (ii) *X* on *Y*, conditional on *R* and *Z.* Similarly, the bias of $\beta_{YX|R}$ and $\beta_{YX|R,Z}$ were estimated empirically by calculating the difference in the *X* coefficient when fitting a linear regression of (i) *Y* on *X*, (ii) *Y* on *X*, conditional on *R*, and (iii) *Y* on *X*, conditional on *R* and *Z*.

The median difference between the theoretical and empirical values of $\beta_{XY|R}$ and $\beta_{XY|R,Z}$ was 0.000 ($5^{th}$ – $95^{th}$ percentile: -0.002 - 0.002) and 0.000 ($5^{th}$ – $95^{th}$ percentile: -0.002 - 0.003), respectively. The median value of the bias of $\beta_{YX|R}$ and $\beta_{YX|R,Z}$ was 0.000 ($5^{th}$ – $95^{th}$ percentile: -0.008 - 0.007) and 0.000 ($5^{th}$ – $95^{th}$ percentile: -0.010 - 0.010), respectively. Therefore, we conclude that the theoretical expressions are correct.

*Section S4. Description of simulation studies to assess the additional bias of the MI estimator from including a predictor of missingness but not the missing values in the imputation model when a binary outcome Y, or continuous or binary exposure X, is partially observed.*

We performed simulation studies to assess the additional bias of the MI estimator from including a predictor of missingness but not the missing values in the imputation model when (i) a binary outcome *Y*, (ii) a continuous exposure *X*, or (iii) a binary exposure *X* were partially observed, in each of the three scenarios discussed in the main paper, considering settings in which we would expect the MI estimator to be biased. For example, in Scenario 1, we considered the setting in which a binary outcome *Y* was partially observed, but not settings in which a continuous or binary exposure *X* was partially observed (because *X* was MAR in Scenario 1 and hence MI using a correctly specified imputation model would be valid).

In each setting, 1000 simulated datasets of size 1000 were generated using the data generating mechanisms described below. We used moderate values of the direct effect sizes (relative to the error variances, which were all equal to one), with all direct effect sizes set to 0.00, 0.50, or 1.00 and the mean of each variable equal to zero. We then set 50% of values of the partially observed variable to missing (in each case, by setting values to missing if $R > 0$). Additional bias was calculated as the average of the per-simulation estimates (where the per-simulation estimate was calculated as the difference between the MI estimate using *Z* and the other analysis model variable as predictors, and the MI estimate using just the other analysis model variable as a predictor in the imputation model, with five imputations in each setting).

*Data generating mechanisms*

In setting (i) (binary outcome *Y* is partially observed), all variables except *Y* were related as defined in Sections S1, S3, and S5 (for Scenarios 1, 2, and 3, respectively), with *Y* defined as $logit\{P(Y{=}1)\} = \beta_{YX}X$ in Scenarios 1 and 3, and $logit\{P(Y{=}1)\} = \beta_{YX}X + \beta_{YU}U$ in Scenario 2. In setting (ii) (continuous exposure *X* is partially observed), all variables were related as defined in Sections S3 and S5 (for Scenarios 2 and 3, respectively). In setting (iii) (binary exposure *X* is partially observed), all variables except *X* were related as defined in Sections S3 and S5 (for Scenarios 2 and 3, respectively), with *X* defined as binary, with probability 0.5 of a value of 0 or 1.

*Results*

Additional bias (and also total bias in Scenario 2) of the MI estimate of $\beta_{YX}$ when the imputation model includes a predictor of missingness, *Z*, when 50% of values are missing for either a binary outcome *Y* (Figures S1, S2-3, and S5 for Scenarios 1-3, respectively), or a binary exposure *X* (Figures S4 and S6 for Scenarios 2 and 3, respectively) are shown below (with results for a continuous exposure *X* illustrated in Figures 4 and 7 in the main text). Note that in these plots, additional and total bias do not take their maximum values because the proportion of missing data is 50% (rather than tending to 100%). However, as per Curnow *et al.* (https://doi.org/10.3389/fepid.2023.1237447), the maximum values are likely to be approximately double the magnitude depicted in these plots.

*Figure S1. Scenario 1. Additional bias of the MI estimate of $\beta_{YX}$ when binary outcome Y is missing not at random, with missingness caused by Y itself, when the imputation model includes an auxiliary variable Z that predicts missingness but not the missing values. Results shown when 50% of values are missing, varying the direct effect sizes $\beta_{YX}$, $\beta_{RY}$, and $\beta_{RZ}$. The distribution of additional bias in each box-plot is due to variation in $\beta_{RY}$.*

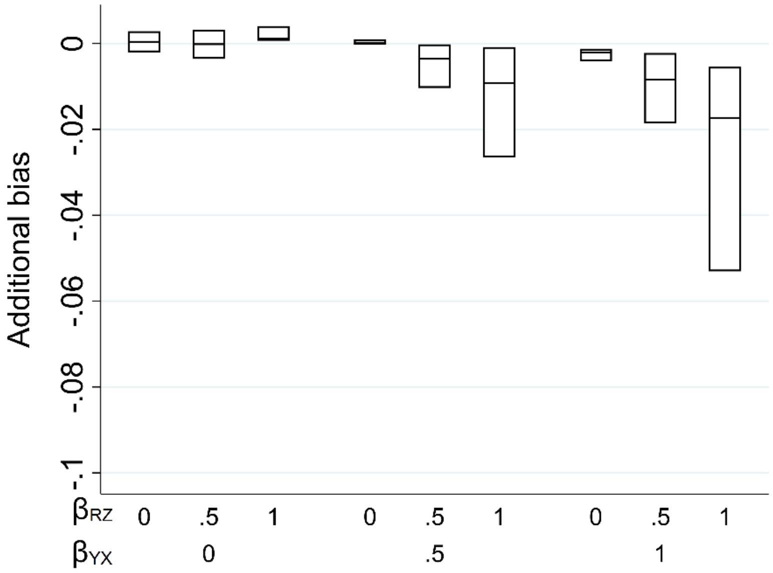

*Figure S2. Scenario 2. Additional bias of the MI estimate of $\beta_{YX}$ when binary outcome Y is missing not at random, with missingness related to Y via an unmeasured variable U, when the imputation model includes an auxiliary variable Z that predicts missingness but not the missing values. Results shown when 50% of values are missing, varying the direct effect sizes $\beta_{YX}$, $\beta_{YU}$, $\beta_{RU}$, and $\beta_{RZ}$. The distribution of additional bias in each box-plot is averaged over the values of $\beta_{YU}$ and $\beta_{RU}$.*

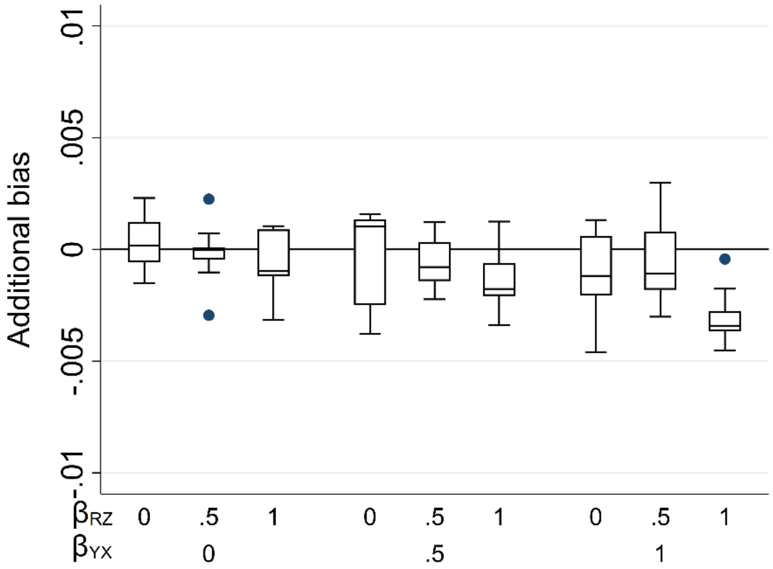


*Figure S3. Scenario 2. Total bias of the MI estimate of $\beta_{YX}$ when binary outcome Y is missing not at random, with missingness related to Y via an unmeasured variable U, when the imputation model includes an auxiliary variable Z that predicts missingness but not the missing values. Results shown when 50% of values are missing, varying the direct effect sizes $\beta_{YX}$, $\beta_{YU}$, $\beta_{RU}$, and $\beta_{RZ}$. The distribution of additional bias in each box-plot is averaged over the values of $\beta_{YU}$ and $\beta_{RU}$.*

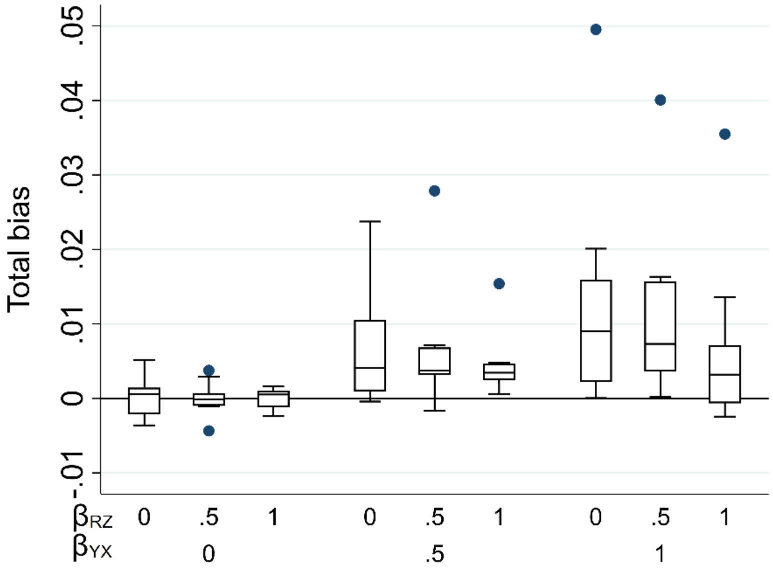

*Figure S4. Scenario 2. Additional bias of the MI estimate of $\beta_{YX}$ when binary exposure X is missing not at random, with missingness related to Y via an unmeasured variable U, when the imputation model includes an auxiliary variable Z that predicts missingness but not the missing values. Results shown when 50% of values are missing, varying the direct effect sizes $\beta_{YX}$, $\beta_{YU}$, $\beta_{RU}$,* and *$\beta_{RZ}$. The distribution of additional bias in each box-plot is averaged over the values of $\beta_{YU}$* and *$\beta_{RU}$.*

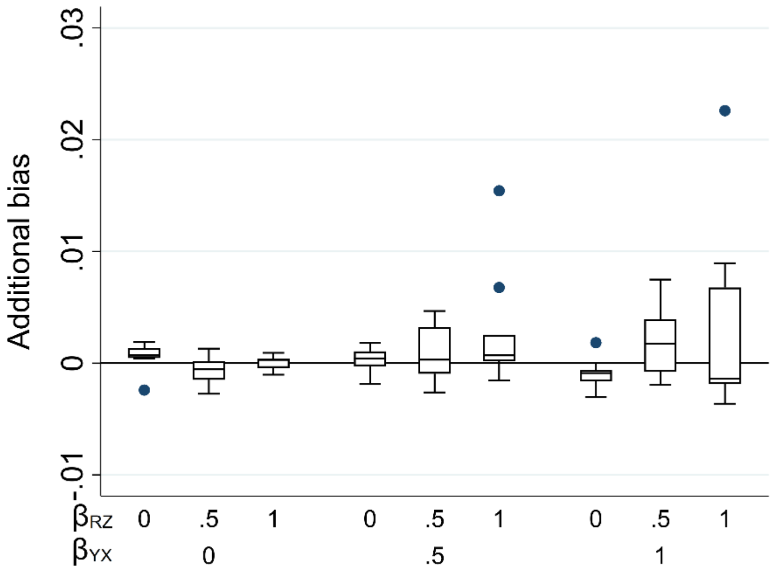


*Figure S5. Scenario 3. Additional bias of the MI estimate of $\beta_{YX}$ when binary outcome Y is missing not at random, with missingness caused by Y and X, when the imputation model includes an auxiliary variable Z that predicts missingness but not the missing values. Results shown when 50% of values are missing, varying the direct effect sizes $\beta_{YX}$, $\beta_{RY}$, $\beta_{RX}$,* and *$\beta_{RZ}$. The distribution of additional bias in each box-plot is averaged over the values of $\beta_{RY}$ and $\beta_{RX}$.*

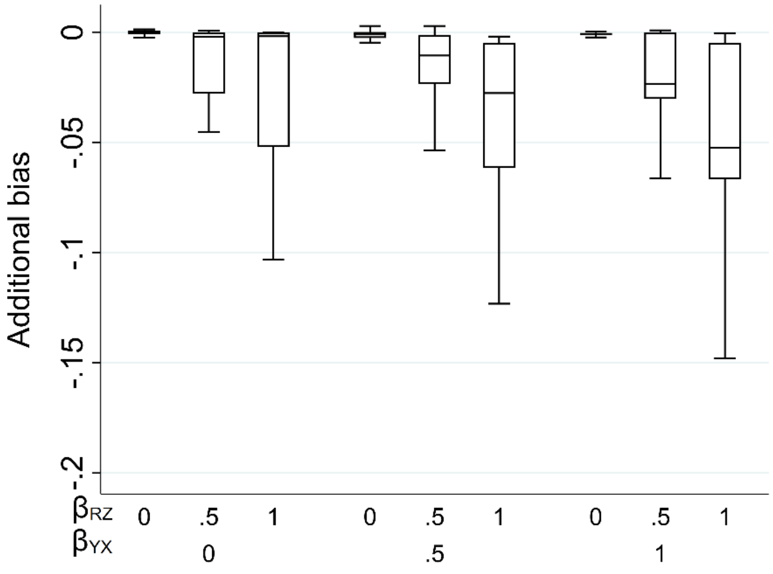

*Figure S6. Scenario 3. Additional bias of the MI estimate of $\beta_{YX}$ when binary exposure X is missing not at random, with missingness caused by Y and X, when the imputation model includes an auxiliary variable Z that predicts missingness but not the missing values. Results shown when 50% of values are missing, varying the direct effect sizes $\beta_{YX}$, $\beta_{RY}$, $\beta_{RX}$,* and *$\beta_{RZ}$. The distribution of additional bias in each box-plot is averaged over the values of $\beta_{RY}$ and $\beta_{RX}$.*

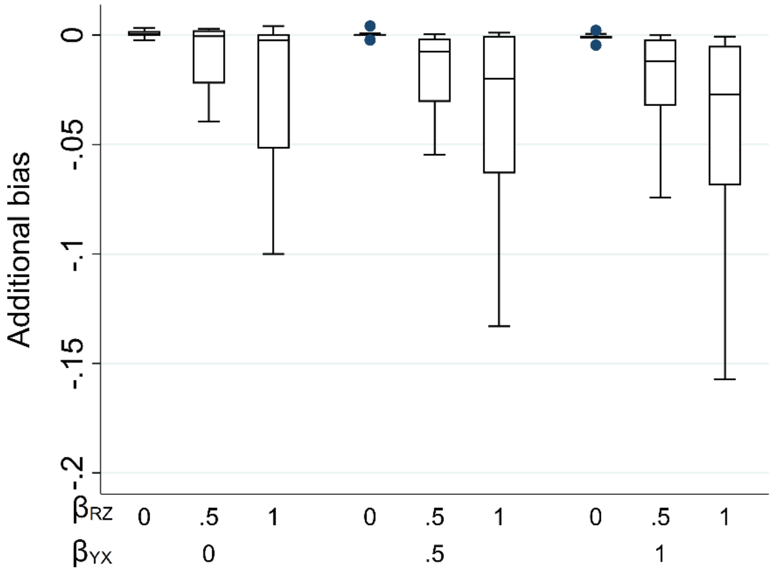

*Section S5. Scenario 3. Derivation of the theoretical expression for the maximum additional bias when a continuous outcome Y or a continuous exposure X is partially observed, with missingness caused by Y and X, when the imputation model includes an auxiliary variable Z that predicts missingness but not the missing values*

As per Figure 4 in the main text, we assume that *Y*, *X*, *Z*, and *R* are normally distributed, such that $Y = \beta_{YX}X + \varepsilon_Y$, where $\varepsilon_Y \sim N(0, \sigma_Y^2)$, $X \sim N(\mu_X, \sigma_X^2)$, $Z \sim N(\mu_Z, \sigma_Z^2)$, and $R = \beta_{RY}Y + \beta_{RX}X + \beta_{RZ}Z + \varepsilon_R$, where $\varepsilon_R \sim N(0, \sigma_R^2)$. We once again assume that each of *Y* and *R* is a linear combination of the variables causing it plus an error term (with *X* and *Z* having no direct causes), with no interactions, all errors uncorrelated, no model mis-specification, and no measurement error, and that an ordinary least squares (OLS) estimator is used to obtain estimates in both analysis and imputation models.

As before, using standard results, the joint distribution of *Y*, *X*, *Z*, and *R*, *f*(*Y*, *X*, *Z*, R), is multivariate normal with covariance matrix ∑, where:

$$\Sigma = \begin{pmatrix} \beta_{YX}^2\sigma_X^2 + \sigma_Y^2 & \beta_{YX}\sigma_X^2 & 0 & \beta_{RY}(\beta_{YX}^2\sigma_X^2 + \sigma_Y^2) + \beta_{YX}\beta_{RX}\sigma_X^2 \\ \beta_{YX}\sigma_X^2 & \sigma_X^2 & 0 & (\beta_{YX}\beta_{RY} + \beta_{RX})\sigma_X^2 \\ 0 & 0 & \sigma_Z^2 & \beta_{RZ}\sigma_Z^2 \\ \beta_{RY}(\beta_{YX}^2\sigma_X^2 + \sigma_Y^2) + \beta_{YX}\beta_{RX}\sigma_X^2 & (\beta_{YX}\beta_{RY} + \beta_{RX})\sigma_X^2 & \beta_{RZ}\sigma_Z^2 & Var(R) \end{pmatrix}$$

where $Var(R) = \beta_{RY}^2(\beta_{YX}^2\sigma_X^2 + \sigma_Y^2) + 2\beta_{YX}\beta_{RY}\beta_{RX}\sigma_X^2 + \beta_{RX}^2\sigma_X^2 + \beta_{RZ}^2\sigma_Z^2 + \sigma_R^2$

Then, the joint conditional distribution of *Y*, *X*, and *Z*, given *R*, *f*(*Y*, *X*, *Z* | *R*=r), is multivariate normal with covariance matrix ∑*, where:

$$\Sigma^* = \begin{pmatrix} \beta_{YX}^2\sigma_X^2 + \sigma_Y^2 & \beta_{YX}\sigma_X^2 & 0 \\ \beta_{YX}\sigma_X^2 & \sigma_X^2 & 0 \\ 0 & 0 & \sigma_Z^2 \end{pmatrix} - \frac{1}{Var(R)} \times$$

$$\begin{pmatrix} \beta_{RY}(\beta_{YX}^2\sigma_X^2 + \sigma_Y^2) + \beta_{YX}\beta_{RX}\sigma_X^2 \\ (\beta_{YX}\beta_{RY} + \beta_{RX})\sigma_X^2 \\ \beta_{RZ}\sigma_Z^2 \end{pmatrix} \begin{pmatrix} \beta_{RY}(\beta_{YX}^2\sigma_X^2 + \sigma_Y^2) + \beta_{YX}\beta_{RX}\sigma_X^2 & (\beta_{YX}\beta_{RY} + \beta_{RX})\sigma_X^2 & \beta_{RZ}\sigma_Z^2 \end{pmatrix}$$

$$= \begin{pmatrix} \beta_{YX}^2\sigma_X^2 + \sigma_Y^2 & \beta_{YX}\sigma_X^2 & 0 \\ \beta_{YX}\sigma_X^2 & \sigma_X^2 & 0 \\ 0 & 0 & \sigma_Z^2 \end{pmatrix} - \frac{1}{Var(R)} \times$$

$$\begin{pmatrix} \{\beta_{RY}(\beta_{YX}^2\sigma_X^2 + \sigma_Y^2) + \beta_{YX}\beta_{RX}\sigma_X^2\}^2 & \dots & \dots \\ (\beta_{YX}\beta_{RY} + \beta_{RX})\{\beta_{RY}(\beta_{YX}^2\sigma_X^2 + \sigma_Y^2) + \beta_{YX}\beta_{RX}\sigma_X^2\}\sigma_X^2 & (\beta_{YX}\beta_{RY} + \beta_{RX})^2\sigma_X^4 & \dots \\ \{\beta_{RY}(\beta_{YX}^2\sigma_X^2 + \sigma_Y^2) + \beta_{YX}\beta_{RX}\sigma_X^2\}\beta_{RZ}\sigma_Z^2 & (\beta_{YX}\beta_{RY} + \beta_{RX})\beta_{RZ}\sigma_X^2\sigma_Z^2 & \beta_{RZ}^2\sigma_Z^4 \end{pmatrix}$$

and the joint conditional distribution of *Y* and *X*, given *Z* and *R*, *f*(*Y*, *X* | *Z*=z, *R*=r), is multivariate normal with covariance matrix ∑**, where:

$$\Sigma^{**} = \begin{pmatrix} \beta_{YX}^2\sigma_X^2 + \sigma_Y^2 & \beta_{YX}\sigma_X^2 \\ \beta_{YX}\sigma_X^2 & \sigma_X^2 \end{pmatrix} - \frac{1}{\sigma_Z^2 Var(R) - \beta_{RZ}^2\sigma_Z^4} \times$$

$$\begin{pmatrix} 0 & \beta_{RY}(\beta_{YX}^2\sigma_X^2 + \sigma_Y^2) + \beta_{YX}\beta_{RX}\sigma_X^2 \\ 0 & (\beta_{YX}\beta_{RY} + \beta_{RX})\sigma_X^2 \end{pmatrix} \begin{pmatrix} Var(R) & -\beta_{RZ}\sigma_Z^2 \\ -\beta_{RZ}\sigma_Z^2 & \sigma_Z^2 \end{pmatrix} \begin{pmatrix} 0 & 0 \\ \beta_{RY}(\beta_{YX}^2\sigma_X^2 + \sigma_Y^2) + \beta_{YX}\beta_{RX}\sigma_X^2 & (\beta_{YX}\beta_{RY} + \beta_{RX})\sigma_X^2 \end{pmatrix}$$

$$= \begin{pmatrix} \beta_{YX}^2\sigma_X^2 + \sigma_Y^2 & \beta_{YX}\sigma_X^2 \\ \beta_{YX}\sigma_X^2 & \sigma_X^2 \end{pmatrix} - \frac{1}{\sigma_Z^2 Var(R) - \beta_{RZ}^2\sigma_Z^4} \times$$

$$\begin{pmatrix} 0 & \beta_{RY}(\beta_{YX}^2\sigma_X^2 + \sigma_Y^2) + \beta_{YX}\beta_{RX}\sigma_X^2 \\ 0 & (\beta_{YX}\beta_{RY} + \beta_{RX})\sigma_X^2 \end{pmatrix} \begin{pmatrix} -\{\beta_{RY}(\beta_{YX}^2\sigma_X^2 + \sigma_Y^2) + \beta_{YX}\beta_{RX}\sigma_X^2\}\beta_{RZ}\sigma_Z^2 & -\beta_{RZ}(\beta_{YX}\beta_{RY} + \beta_{RX})\sigma_X^2\sigma_Z^2 \\ \{\beta_{RY}(\beta_{YX}^2\sigma_X^2 + \sigma_Y^2) + \beta_{YX}\beta_{RX}\sigma_X^2\}\sigma_Z^2 & (\beta_{YX}\beta_{RY} + \beta_{RX})\sigma_X^2\sigma_Z^2 \end{pmatrix}$$

$$= \begin{pmatrix} \beta_{YX}^2\sigma_X^2 + \sigma_Y^2 & \beta_{YX}\sigma_X^2 \\ \beta_{YX}\sigma_X^2 & \sigma_X^2 \end{pmatrix} - \frac{1}{\sigma_Z^2 Var(R) - \beta_{RZ}^2\sigma_Z^4} \times$$

$$\begin{pmatrix} (\beta_{RY}(\beta_{YX}^2\sigma_X^2 + \sigma_Y^2) + \beta_{YX}\beta_{RX}\sigma_X^2)^2\sigma_Z^2 & (\beta_{RY}(\beta_{YX}^2\sigma_X^2 + \sigma_Y^2) + \beta_{YX}\beta_{RX}\sigma_X^2)(\beta_{YX}\beta_{RY} + \beta_{RX})\sigma_X^2\sigma_Z^2 \\ (\beta_{RY}(\beta_{YX}^2\sigma_X^2 + \sigma_Y^2) + \beta_{YX}\beta_{RX}\sigma_X^2)(\beta_{YX}\beta_{RY} + \beta_{RX})\sigma_X^2\sigma_Z^2 & (\beta_{YX}\beta_{RY} + \beta_{RX})^2\sigma_X^4\sigma_Z^2 \end{pmatrix}$$

As per the main text, we consider the bias of the MI estimator in two separate settings where: (i) *Y* is partially observed, and (ii) *X* is partially observed.

**Setting 1: *Y* is partially observed**

Consider the *X* coefficient from the imputation model for *Y* (when only *X* is used as a predictor), which tends to $\beta_{YX|R}$ as the proportion of missing data tends to one.

Using the results above, $\beta_{YX|R} = \frac{Cov(X,Y|R)}{Var(X|R)}$

$$= \frac{\beta_{YX}\sigma_X^2 Var(R) - (\beta_{YX}\beta_{RY} + \beta_{RX})\{\beta_{RY}(\beta_{YX}^2\sigma_X^2 + \sigma_Y^2) + \beta_{YX}\beta_{RX}\sigma_X^2\}\sigma_X^2}{\sigma_X^2 Var(R) - (\beta_{YX}\beta_{RY} + \beta_{RX})^2\sigma_X^4}$$

$$= \frac{\beta_{YX}\{\beta_{RY}^2(\beta_{YX}^2\sigma_X^2 + \sigma_Y^2) + 2\beta_{RY}\beta_{RX}\beta_{YX}\sigma_X^2 + \beta_{RX}^2\sigma_X^2 + \beta_{RZ}^2\sigma_Z^2 + \sigma_R^2\}}{\beta_{RY}^2(\beta_{YX}^2\sigma_X^2 + \sigma_Y^2) + 2\beta_{YX}\beta_{RY}\beta_{RX}\sigma_X^2 + \beta_{RX}^2\sigma_X^2 + \beta_{RZ}^2\sigma_Z^2 + \sigma_R^2 - (\beta_{YX}\beta_{RY} + \beta_{RX})^2\sigma_X^2}$$

$$- \left\{\frac{(\beta_{YX}\beta_{RY} + \beta_{RX})\{\beta_{RY}(\beta_{YX}^2\sigma_X^2 + \sigma_Y^2) + \beta_{YX}\beta_{RX}\sigma_X^2\}}{\beta_{RY}^2(\beta_{YX}^2\sigma_X^2 + \sigma_Y^2) + 2\beta_{YX}\beta_{RY}\beta_{RX}\sigma_X^2 + \beta_{RX}^2\sigma_X^2 + \beta_{RZ}^2\sigma_Z^2 + \sigma_R^2 - (\beta_{YX}\beta_{RY} + \beta_{RX})^2\sigma_X^2}\right\}$$

$$= \frac{\beta_{YX}\beta_{RY}^2(\beta_{YX}^2\sigma_X^2 + \sigma_Y^2) + 2\beta_{YX}\beta_{RY}\beta_{RX}\beta_{YX}\sigma_X^2 + \beta_{YX}\beta_{RX}^2\sigma_X^2 + \beta_{YX}\beta_{RZ}^2\sigma_Z^2 + \beta_{YX}\sigma_R^2}{\beta_{YX}^2\beta_{RY}^2\sigma_X^2 + \beta_{RY}^2\sigma_Y^2 + 2\beta_{YX}\beta_{RY}\beta_{RX}\sigma_X^2 + \beta_{RX}^2\sigma_X^2 + \beta_{RZ}^2\sigma_Z^2 + \sigma_R^2 - \beta_{YX}^2\beta_{RY}^2\sigma_X^2 - \beta_{RX}^2\sigma_X^2 - 2\beta_{YX}\beta_{RY}\beta_{RX}\sigma_X^2}$$

$$- \left\{\frac{\beta_{YX}\beta_{RY}^2(\beta_{YX}^2\sigma_X^2 + \sigma_Y^2) + \beta_{RX}\beta_{RY}(\beta_{YX}^2\sigma_X^2 + \sigma_Y^2) + \beta_{YX}^2\beta_{RY}\beta_{RX}\sigma_X^2 + \beta_{YX}\beta_{RX}^2\sigma_X^2\}}{\beta_{YX}^2\beta_{RY}^2\sigma_X^2 + \beta_{RY}^2\sigma_Y^2 + 2\beta_{YX}\beta_{RY}\beta_{RX}\sigma_X^2 + \beta_{RX}^2\sigma_X^2 + \beta_{RZ}^2\sigma_Z^2 + \sigma_R^2 - \beta_{YX}^2\beta_{RY}^2\sigma_X^2 - \beta_{RX}^2\sigma_X^2 - 2\beta_{YX}\beta_{RY}\beta_{RX}\sigma_X^2}\right\}$$

$$= \frac{\beta_{YX}\beta_{RZ}^2\sigma_Z^2 + \beta_{YX}\sigma_R^2 - \beta_{RY}\beta_{RX}\sigma_Y^2}{\beta_{RY}^2\sigma_Y^2 + \beta_{RZ}^2\sigma_Z^2 + \sigma_R^2}$$

$$= \beta_{YX} \times \left\{\frac{\beta_{RZ}^2\sigma_Z^2 + \sigma_R^2 - \frac{\beta_{RY}\beta_{RX}\sigma_Y^2}{\beta_{YX}}}{\beta_{RY}^2\sigma_Y^2 + \sigma_R^2 + \beta_{RZ}^2\sigma_Z^2}\right\}$$

$$= \beta_{YX} \times \left\{1 - \frac{\left(\beta_{RY} + \frac{\beta_{RX}}{\beta_{YX}}\right)\beta_{RY}\sigma_Y^2}{\beta_{RY}^2\sigma_Y^2 + \sigma_R^2 + \beta_{RZ}^2\sigma_Z^2}\right\}$$

Similarly, the *X* coefficient from the imputation model for *Y* when *X* and *Z* are used as predictors tends to $\beta_{YX|R,Z}$ as the proportion of missing data tends to one, where:

$$\beta_{YX|R,Z} = \frac{Cov(X,Y|R,Z)}{Var(X|R,Z)}$$

$$= \frac{\beta_{YX}\sigma_X^2(\sigma_Z^2 Var(R) - \beta_{RZ}^2\sigma_Z^4) - (\beta_{RY}(\beta_{YX}^2\sigma_X^2 + \sigma_Y^2) + \beta_{YX}\beta_{RX}\sigma_X^2)(\beta_{YX}\beta_{RY} + \beta_{RX})\sigma_X^2\sigma_Z^2}{\sigma_X^2(\sigma_Z^2 Var(R) - \beta_{RZ}^2\sigma_Z^4) - (\beta_{YX}\beta_{RY} + \beta_{RX})^2\sigma_X^4\sigma_Z^2}$$

$$= \frac{\beta_{YX}\sigma_X^2\sigma_Z^2\{\beta_{RY}^2(\beta_{YX}^2\sigma_X^2 + \sigma_Y^2) + 2\beta_{RY}\beta_{RX}\beta_{YX}\sigma_X^2 + \beta_{RX}^2\sigma_X^2 + \beta_{RZ}^2\sigma_Z^2 + \sigma_R^2 - \beta_{RZ}^2\sigma_Z^2\}}{\sigma_X^2\sigma_Z^2(\beta_{RY}^2(\beta_{YX}^2\sigma_X^2 + \sigma_Y^2) + 2\beta_{RY}\beta_{RX}\beta_{YX}\sigma_X^2 + \beta_{RX}^2\sigma_X^2 + \beta_{RZ}^2\sigma_Z^2 + \sigma_R^2 - \beta_{RZ}^2\sigma_Z^2 - (\beta_{YX}\beta_{RY} + \beta_{RX})^2\sigma_X^2)}$$

$$- \left\{\frac{\sigma_X^2\sigma_Z^2(\beta_{RY}(\beta_{YX}^2\sigma_X^2 + \sigma_Y^2) + \beta_{YX}\beta_{RX}\sigma_X^2)(\beta_{YX}\beta_{RY} + \beta_{RX})}{\sigma_X^2\sigma_Z^2(\beta_{RY}^2(\beta_{YX}^2\sigma_X^2 + \sigma_Y^2) + 2\beta_{RY}\beta_{RX}\beta_{YX}\sigma_X^2 + \beta_{RX}^2\sigma_X^2 + \beta_{RZ}^2\sigma_Z^2 + \sigma_R^2 - \beta_{RZ}^2\sigma_Z^2 - (\beta_{YX}\beta_{RY} + \beta_{RX})^2\sigma_X^2)}\right\}$$

$$= \frac{\beta_{YX}\beta_{RY}^2(\beta_{YX}^2\sigma_X^2 + \sigma_Y^2) + 2\beta_{YX}^2\beta_{RY}\beta_{RX}\sigma_X^2 + \beta_{YX}\beta_{RX}^2\sigma_X^2 + \beta_{YX}\sigma_R^2}{\beta_{RY}^2\sigma_Y^2 + \sigma_R^2} - \left\{\frac{\beta_{YX}\beta_{RY}^2(\beta_{YX}^2\sigma_X^2 + \sigma_Y^2) + 2\beta_{YX}^2\beta_{RY}\beta_{RX}\sigma_X^2 + \beta_{YX}\beta_{RX}^2\sigma_X^2 + \beta_{RY}\beta_{RX}\sigma_Y^2}{\beta_{RY}^2\sigma_Y^2 + \sigma_R^2}\right\}$$

$$= \beta_{YX} \times \left\{\frac{\sigma_R^2 - \frac{\beta_{RY}\beta_{RX}\sigma_Y^2}{\beta_{YX}}}{\beta_{RY}^2\sigma_Y^2 + \sigma_R^2}\right\}$$

$$= \beta_{YX} \times \left\{1 - \frac{\beta_{RY}\sigma_Y^2\left(\beta_{RY} + \frac{\beta_{RX}}{\beta_{YX}}\right)}{\beta_{RY}^2\sigma_Y^2 + \sigma_R^2}\right\}$$

Thus, the maximum bias of the MI estimator due to *Y* being MNAR (using only *X* as a predictor in the imputation model for *Y*) is $-\frac{\beta_{YX}\beta_{RY}\sigma_Y^2\left(\beta_{RY}+\frac{\beta_{RX}}{\beta_{YX}}\right)}{\beta_{RY}^2\sigma_Y^2+\sigma_R^2+\beta_{RZ}^2\sigma_Z^2}$. The maximum additional bias of the MI estimator (*i.e.* in addition to the bias due to *Y* being MNAR) from including *Z* as a predictor in the imputation model is $\beta_{YX}\beta_{RY}\sigma_Y^2\left(\beta_{RY} + \frac{\beta_{RX}}{\beta_{YX}}\right) \times \left\{\frac{1}{\beta_{RY}^2\sigma_Y^2+\sigma_R^2+\beta_{RZ}^2\sigma_Z^2} - \frac{1}{\beta_{RY}^2\sigma_Y^2+\sigma_R^2}\right\} = \frac{-\beta_{YX}\beta_{RY}\beta_{RZ}^2\sigma_Y^2\sigma_Z^2\left(\beta_{RY}+\frac{\beta_{RX}}{\beta_{YX}}\right)}{(\beta_{RY}^2\sigma_Y^2+\sigma_R^2+\beta_{RZ}^2\sigma_Z^2)(\beta_{RY}^2\sigma_Y^2+\sigma_R^2)}$ as per Formula 4.2 in the main text.

Or in other words, if bias amplification is defined as the bias of $\beta_{YX|Z,R}$ divided by the bias of $\beta_{YX|R}$, then the maximum bias amplification is: $\frac{-\beta_{YX}\beta_{RY}\sigma_Y^2\left(\beta_{RY}+\frac{\beta_{RX}}{\beta_{YX}}\right)}{\beta_{RY}^2\sigma_Y^2+\sigma_R^2} / \frac{-\beta_{YX}\beta_{RY}\sigma_Y^2\left(\beta_{RY}+\frac{\beta_{RX}}{\beta_{YX}}\right)}{\beta_{RZ}^2\sigma_Z^2+\beta_{RY}^2\sigma_Y^2+\sigma_R^2} =$

$\frac{\beta_{RZ}^2\sigma_Z^2+\beta_{RY}^2\sigma_Y^2+\sigma_R^2}{\beta_{RY}^2\sigma_Y^2+\sigma_R^2} = 1 + \frac{\beta_{RZ}^2\sigma_Z^2}{\beta_{RY}^2\sigma_Y^2+\sigma_R^2}$, that is, the maximum bias due to *Y* being MNAR is amplified by a factor of $\left\{1 + \frac{\beta_{RZ}^2\sigma_Z^2}{\beta_{RY}^2\sigma_Y^2+\sigma_R^2}\right\}$ when *Z* is included in the imputation model for *Y*.

**Setting 2: *X* is partially observed**

Consider the *Y* coefficient from the imputation model for *X* (when only *Y* is used as a predictor), which tends to $\beta_{XY|R}$ as the proportion of missing data tends to one.

$$\beta_{XY|R} = \frac{Cov(X,Y|R)}{Var(Y|R)}$$

$$= \frac{\beta_{YX}\sigma_X^2 Var(R) - (\beta_{YX}\beta_{RY} + \beta_{RX})\{\beta_{RY}(\beta_{YX}^2\sigma_X^2 + \sigma_Y^2) + \beta_{YX}\beta_{RX}\sigma_X^2\}\sigma_X^2}{(\beta_{YX}^2\sigma_X^2 + \sigma_Y^2)Var(R) - \{\beta_{RY}(\beta_{YX}^2\sigma_X^2 + \sigma_Y^2) + \beta_{YX}\beta_{RX}\sigma_X^2\}^2}$$

Using results above and noting $\{\beta_{RY}(\beta_{YX}^2\sigma_X^2 + \sigma_Y^2) + \beta_{YX}\beta_{RX}\sigma_X^2\}^2 = \beta_{RY}^2(\beta_{YX}^2\sigma_X^2 + \sigma_Y^2)^2 + \beta_{YX}^2\beta_{RX}^2\sigma_X^4 + 2\beta_{YX}\beta_{RY}\beta_{RX}\sigma_X^2(\beta_{YX}^2\sigma_X^2 + \sigma_Y^2)$, this expression can be simplified to:

$$\beta_{XY|R} = \frac{\beta_{YX}\sigma_X^2\{\beta_{RZ}^2\sigma_Z^2 + \sigma_R^2 - \frac{\beta_{RY}\beta_{RX}\sigma_Y^2}{\beta_{YX}}\}}{(\beta_{YX}^2\sigma_X^2 + \sigma_Y^2)(\beta_{RX}^2\sigma_X^2 + \beta_{RZ}^2\sigma_Z^2 + \sigma_R^2) - \beta_{YX}^2\beta_{RX}^2\sigma_X^4}$$

$$= \frac{\beta_{YX}\sigma_X^2\{\beta_{RZ}^2\sigma_Z^2 + \sigma_R^2 - \frac{\beta_{RY}\beta_{RX}\sigma_Y^2}{\beta_{YX}}\}}{(\beta_{YX}^2\sigma_X^2 + \sigma_Y^2)(\beta_{RX}^2\sigma_X^2 + \beta_{RZ}^2\sigma_Z^2 + \sigma_R^2) - \beta_{YX}^2\beta_{RX}^2\sigma_X^4}$$

$$= \frac{\beta_{YX}\sigma_X^2}{(\beta_{YX}^2\sigma_X^2 + \sigma_Y^2)} \times \frac{\beta_{RZ}^2\sigma_Z^2 + \sigma_R^2 - \frac{\beta_{RY}\beta_{RX}\sigma_Y^2}{\beta_{YX}}}{\beta_{RX}^2\sigma_X^2 + \beta_{RZ}^2\sigma_Z^2 + \sigma_R^2 - \{\beta_{YX}^2\beta_{RX}^2\sigma_X^4/(\beta_{YX}^2\sigma_X^2 + \sigma_Y^2)\}}$$

$$= \beta_{XY} \times \left\{1 - \frac{\beta_{RX}\left\{\frac{\beta_{RY}\sigma_Y^2}{\beta_{YX}} + \beta_{RX}\sigma_X^2\left(1 - \frac{\beta_{YX}^2\sigma_X^2}{\beta_{YX}^2\sigma_X^2 + \sigma_Y^2}\right)\right\}}{\beta_{RX}^2\sigma_X^2 + \beta_{RZ}^2\sigma_Z^2 + \sigma_R^2 - \{\beta_{YX}^2\beta_{RX}^2\sigma_X^4/(\beta_{YX}^2\sigma_X^2 + \sigma_Y^2)\}}\right\}$$

Similarly, the *Y* coefficient from the imputation model for *X* when *Y* and *Z* are used as predictors tends to $\beta_{XY|R,Z}$ as the proportion of missing data tends to one.

$$\beta_{XY|R,Z} = \frac{Cov(X,Y|R,Z)}{Var(Y|R,Z)}$$

$$= \frac{\beta_{YX}\sigma_X^2(\sigma_Z^2 Var(R) - \beta_{RZ}^2\sigma_Z^4) - (\beta_{RY}(\beta_{YX}^2\sigma_X^2 + \sigma_Y^2) + \beta_{YX}\beta_{RX}\sigma_X^2)(\beta_{YX}\beta_{RY} + \beta_{RX})\sigma_X^2\sigma_Z^2}{(\beta_{YX}^2\sigma_X^2 + \sigma_Y^2)(\sigma_Z^2 Var(R) - \beta_{RZ}^2\sigma_Z^4) - \{\beta_{RY}(\beta_{YX}^2\sigma_X^2 + \sigma_Y^2) + \beta_{YX}\beta_{RX}\sigma_X^2\}^2\sigma_Z^2}$$

Using results above, this expression can be simplified to:

$$\beta_{XY|R,Z} = \frac{\beta_{YX}\sigma_X^2\{\sigma_R^2 - \frac{\beta_{RY}\beta_{RX}\sigma_Y^2}{\beta_{YX}}\}}{(\beta_{YX}^2\sigma_X^2 + \sigma_Y^2)(\beta_{RX}^2\sigma_X^2 + \sigma_R^2) - \beta_{YX}^2\beta_{RX}^2\sigma_X^4}$$

$$= \frac{\beta_{YX}\sigma_X^2}{(\beta_{YX}^2\sigma_X^2 + \sigma_Y^2)} \times \frac{\sigma_R^2 - \frac{\beta_{RY}\beta_{RX}\sigma_Y^2}{\beta_{YX}}}{\beta_{RX}^2\sigma_X^2 + \sigma_R^2 - \{\beta_{YX}^2\beta_{RX}^2\sigma_X^4/(\beta_{YX}^2\sigma_X^2 + \sigma_Y^2)\}}$$

$$= \beta_{XY} \times \left\{ 1 - \frac{\frac{\beta_{RY}\beta_{RX}\sigma_Y^2}{\beta_{YX}} + \beta_{RX}^2\sigma_X^2 - \left\{\frac{\beta_{YX}^2\beta_{RX}^2\sigma_X^4}{(\beta_{YX}^2\sigma_X^2 + \sigma_Y^2)}\right\}}{\beta_{RX}^2\sigma_X^2 + \sigma_R^2 - \{\beta_{YX}^2\beta_{RX}^2\sigma_X^4/(\beta_{YX}^2\sigma_X^2 + \sigma_Y^2)\}} \right\}$$

$$= \beta_{XY} \times \left\{ 1 - \frac{\beta_{RX}\left\{\frac{\beta_{RY}\sigma_Y^2}{\beta_{YX}} + \beta_{RX}\sigma_X^2\left(1 - \frac{\beta_{YX}^2\sigma_X^2}{\beta_{YX}^2\sigma_X^2 + \sigma_Y^2}\right)\right\}}{\beta_{RX}^2\sigma_X^2 + \sigma_R^2 - \{\beta_{YX}^2\beta_{RX}^2\sigma_X^4/(\beta_{YX}^2\sigma_X^2 + \sigma_Y^2)\}} \right\}$$

Thus, the maximum additional bias of the *Y* coefficient in the imputation model for *X* (*i.e.* in addition to the bias due to *X* being MNAR) from including *Z* as a predictor in the imputation model is: $\beta_{XY}\beta_{RX}\left\{\frac{\beta_{RY}\sigma_Y^2}{\beta_{YX}} + \beta_{RX}\sigma_X^2\left(1 - \frac{\beta_{YX}^2\sigma_X^2}{\beta_{YX}^2\sigma_X^2+\sigma_Y^2}\right)\right\} \times \left\{\frac{1}{\beta_{RX}^2\sigma_X^2+\beta_{RZ}^2\sigma_Z^2+\sigma_R^2-\{\beta_{YX}^2\beta_{RX}^2\sigma_X^4/(\beta_{YX}^2\sigma_X^2+\sigma_Y^2)\}} - \frac{1}{\beta_{RX}^2\sigma_X^2+\sigma_R^2-\{\beta_{YX}^2\beta_{RX}^2\sigma_X^4/(\beta_{YX}^2\sigma_X^2+\sigma_Y^2)\}}\right\}$ as per Formula 4.4 in the main text.

As before, the theoretical expressions for the maximum bias of $\beta_{YX|R}$ and $\beta_{XY|R}$, as well as the maximum additional bias of $\beta_{YX|R,Z}$ and $\beta_{XY|R,Z}$, were verified using simulation. We used 1000 simulations, and each simulated dataset contained 100,000 observations. In each simulated dataset, the values of each coefficient ($\beta_{YX}$, $\beta_{RZ}$, *etc.*) and each error variance ($\sigma_X^2$, $\sigma_Z^2$, *etc.*) were sampled from a uniform distribution *U*(0, 2). For simplicity, $\mu_X$ and $\mu_Z$ were set equal to zero (note that the formulas do not depend on these parameters). Data were then generated using the models for *Y*, *X*, *Z*, and *R* that were specified above. All bias quantities were calculated using the theoretical expressions. They were also estimated empirically by calculating (for Setting 1) the difference in the *X* coefficient when fitting a linear regression of (i) *Y* on *X*, (ii) *Y* on *X*, conditional on *R*, and (iii) *Y* on *X*, conditional on *R* and *Z*, and (for Setting 2) the difference in the *Y* coefficient when fitting a linear regression of (i) *X* on *Y*, (ii) *X* on *Y*, conditional on *R*, and (iii) *X* on *Y*, conditional on *R* and *Z* (with, in each setting, the difference between the coefficient from models (i) and (ii) used to estimate the maximum bias, and the difference between the coefficient from models (ii) and (iii) used to estimate the maximum additional bias).

In Setting 1, the median difference between the theoretical and empirical values of maximum bias and maximum additional bias was 0.000 ($5^{th}$ – $95^{th}$ percentile: -0.008 - 0.009) and 0.000 ($5^{th}$ – $95^{th}$ percentile: -0.006 - 0.006), respectively. In Setting 2, the median difference between the theoretical and empirical values of maximum bias and maximum additional bias was 0.000 ($5^{th}$ – $95^{th}$ percentile: -0.004 - 0.003) and 0.000 ($5^{th}$ – $95^{th}$ percentile: -0.003 - 0.003), respectively. Therefore, we conclude that all theoretical expressions are correct.

*Section S6. Stata code for formula verification and illustration, and to generate data as per the simulation studies*

```
*** Scenario 1  ***
* 1. Verification of bias

*Define postfile to store results
tempname simloop
postfile `simloop' int(i) float(b_yx beta_yx beta_yx_cond_r
beta_yx_cond_rz theor_bias emp_bias theor_addbias emp_addbias) using
"sim_yxzr_scen1.dta", replace

forvalues i=1/1000 {
      clear
      *SD
      local s_z=runiform(0,2)
      local s_r=runiform(0,2)
      local s_x=runiform(0,2)
      local s_y=runiform(0,2)
      *beta
      local b_yx=runiform(0,2)
      local b_ry=runiform(0,2)
      local b_rz=runiform(0,2)

      *RVs
      quietly set obs 100000
      gen z=rnormal(0,`s_z')
      gen x=rnormal(0,`s_x')
      gen y=rnormal(`b_yx'*x,`s_y')
      gen r=rnormal(`b_ry'*y + `b_rz'*z,`s_r')

      *Estimate beta_YX
      quietly regress y x
      local beta_yx=e(b)[1,1]

      *Estimate beta_YX|R
      quietly regress y x r
      *Store estimate
      local beta_yx_cond_r=e(b)[1,1]

      *Estimate beta_YX|R,Z
      quietly regress y x r z
      *Store estimate
      local beta_yx_cond_rz=e(b)[1,1]

      *Calculate theoretical and empirical bias and bias amp
      local emp_bias = `beta_yx_cond_r' - `beta_yx'
      local emp_addbias = `beta_yx_cond_rz' - `beta_yx_cond_r'

      local theor_bias=-`b_yx'*`b_ry'^2*`s_y'^2/(`b_ry'^2*`s_y'^2 +
`s_r'^2 + `b_rz'^2*`s_z'^2)
      local
theor_addbias=`b_yx'*`b_ry'^2*`s_y'^2*((1/(`b_ry'^2*`s_y'^2 +
`s_r'^2 + `b_rz'^2*`s_z'^2)) - (1/(`b_ry'^2*`s_y'^2 + `s_r'^2)))
```

```
post `simloop' (`i') (`b_yx') (`beta_yx') (`beta_yx_cond_r')
(`beta_yx_cond_rz') (`theor_bias') (`emp_bias') (`theor_addbias')
(`emp_addbias')
}
postclose `simloop'

use sim_yxzr_scen1, clear
gen diff_bias=theor_bias - emp_bias
gen diff_addbias=theor_addbias-emp_addbias
sum diff_bias diff_addbias, detail

*2. Illustration of max additional bias formula
****** Varying the size of parameters - still use mu_z=0 and mu_u=0
and all error vars = 1*******

*Define postfile to store results
postfile `simloop' float(b_yx b_ry b_rz max_addbias max_totalbias
max_biasmnar) using "maxaddbias_illustration_scen1.dta", replace

foreach b_yx of numlist 0 0.25 0.5 0.75 1 {
      foreach b_ry of numlist 0 0.25 0.5 0.75 1 {
            foreach b_rz of numlist 0 0.25 0.5 0.75 1 {
                              clear
                              local
max_addbias=`b_yx'*`b_ry'^2*(1/(`b_ry'^2 + `b_rz'^2 + 1) -
1/(`b_ry'^2 + 1))
                              local max_totalbias=-
`b_yx'*`b_ry'^2/(`b_ry'^2 + 1)
                              local max_biasmnar=-
`b_yx'*`b_ry'^2/(`b_ry'^2 + `b_rz'^2 + 1)

post `simloop' (`b_yx') (`b_ry') (`b_rz') (`max_addbias')
(`max_totalbias') (`max_biasmnar')
            }
      }
}
postclose `simloop'


* 3. Illustration of max additional bias - Binary Y
****** Varying the size of parameters - still use mu_z=0 and mu_u=0
and all error vars = 1*******
tempname simloop
postfile `simloop' int(nsim) float(b_yx b_ry b_rz beta_x_x se_x_x
beta_x_xz se_x_xz) using "MIaddbias_illustration_scen1Ybin.dta",
replace

*Create a temporary file for storing simulated data
tempfile tmpfull

forvalues nsim=1/1000 {
      *print i
      di "`nsim'"

      foreach b_yx of numlist 0 0.5 1 {
            foreach b_ry of numlist 0 0.5 1 {
```

```
                foreach b_rz of numlist 0 0.5 1 {
                        clear
                        quietly set obs 1000
                        gen z=rnormal(0,1)
                        gen x=rnormal(0,1)
                        gen y=rbinomial(1,invlogit(`b_yx'*x ))
                        gen r=rnormal(`b_ry'*y + `b_rz'*z,1)

                        gen ymiss=y
                        quietly replace ymiss=. if r>0
                        quietly save `tmpfull', replace

                        *MI with X
                        quietly mi set flong
                        quietly mi register imputed ymiss
                        quietly mi register regular x
                        quietly mi impute logit ymiss = x, add(5)
                        quietly mi estimate: logistic ymiss x
                        local beta_x = e(b_mi)[1,1]
                        local se_x = sqrt(e(V_mi)[1,1])

                        *MI with X and Z
                    *restore simulated data i.e. before imputation
                    use `tmpfull', clear
                    quietly mi set flong
                    quietly mi register imputed ymiss
                    quietly mi register regular x z
                    quietly mi impute logit ymiss = x z, add(5)
                    quietly mi estimate: logistic ymiss x

post `simloop' (`nsim') (`b_yx') (`b_ry') (`b_rz') (`beta_x')
(`se_x') (e(b_mi)[1,1]) (sqrt(e(V_mi)[1,1]))
                }
            }
        }
}
postclose `simloop'


*** Scenario 2  ***
* 1. Verification of bias

*Define postfile to store results
tempname simloop
postfile `simloop' int(i) float(b_yx beta_yx beta_yx_cond_r
beta_yx_cond_rz beta_xy_cond_r beta_xy_cond_rz theor_beta_xy_cond_r
theor_beta_xy_cond_rz) using "sim_yxzru_scen2.dta", replace

forvalues i=1/1000 {
    *di "`i'"
    clear
    *SD
    local s_z=runiform(0,2)
    local s_r=runiform(0,2)
    local s_x=runiform(0,2)
```

```
      local s_y=runiform(0,2)
      local s_u=runiform(0,2)
      *beta
      local b_yx=runiform(0,2)
      local b_yu=runiform(0,2)
      local b_rz=runiform(0,2)
      local b_ru=runiform(0,2)

      *RVs
      quietly set obs 100000
      gen z=rnormal(0,`s_z')
      gen x=rnormal(0,`s_x')
      gen u=rnormal(0,`s_u')
      gen y=rnormal(`b_yx'*x + `b_yu'*u,`s_y')
      gen r=rnormal(`b_ru'*u + `b_rz'*z,`s_r')

      *Estimate beta_YX
      quietly regress y x
      local beta_yx=e(b)[1,1]

      *Estimate beta_YX|R
      quietly regress y x r
      *Store estimate
      local beta_yx_cond_r=e(b)[1,1]

      *Estimate beta_YX|R,Z
      quietly regress y x r z
      *Store estimate
      local beta_yx_cond_rz=e(b)[1,1]

      *Estimate beta_XY|R
      quietly regress x y r
      *Store estimate
      local beta_xy_cond_r=e(b)[1,1]

      *Estimate beta_XY|R,Z
      quietly regress x y r z
      *Store estimate
      local beta_xy_cond_rz=e(b)[1,1]

      *Calculate theoretical values
      local theor_beta_xy_cond_r =(`b_yx'*`s_x'^2/(`b_yx'^2*`s_x'^2
+ `b_yu'^2*`s_u'^2 + `s_y'^2)) * 1/(1-
(`b_yu'^2*`b_ru'^2*`s_u'^4/((`b_yx'^2*`s_x'^2 + `b_yu'^2*`s_u'^2 +
`s_y'^2)*(`b_rz'^2*`s_z'^2 + `b_ru'^2*`s_u'^2 + `s_r'^2))))
      local theor_beta_xy_cond_rz =(`b_yx'*`s_x'^2/(`b_yx'^2*`s_x'^2
+ `b_yu'^2*`s_u'^2 + `s_y'^2)) * 1/(1-
(`b_yu'^2*`b_ru'^2*`s_u'^4/((`b_yx'^2*`s_x'^2 + `b_yu'^2*`s_u'^2 +
`s_y'^2)*(`b_ru'^2*`s_u'^2 + `s_r'^2))))

post `simloop' (`i') (`b_yx') (`beta_yx') (`beta_yx_cond_r')
(`beta_yx_cond_rz') (`beta_xy_cond_r') (`beta_xy_cond_rz')
(`theor_beta_xy_cond_r') (`theor_beta_xy_cond_rz')
}
postclose `simloop'
```

```
use sim_yxzru_scen2, clear
gen bias_beta_yx_cond_r=beta_yx_cond_r-beta_yx
gen bias_beta_yx_cond_rz=beta_yx_cond_rz-beta_yx
gen diff_beta_xy_cond_r=theor_beta_xy_cond_r-beta_xy_cond_r
gen diff_beta_xy_cond_rz=theor_beta_xy_cond_rz-beta_xy_cond_rz

sum bias_beta_yx_cond_r bias_beta_yx_cond_rz diff_beta_xy_cond_r
diff_beta_xy_cond_rz, detail

* 2. Additional bias – X continuous
****** Varying the size of parameters - still use mu_z=0 and mu_u=0
and all error vars = 1*******
tempname simloop
postfile `simloop' int(nsim) float(b_yx b_yu b_ru b_rz beta_x_y
se_x_y beta_x_yz se_x_yz) using
"MIaddbias_illustration_scen2Xcts.dta", replace

*Create a temporary file for storing simulated data
tempfile tmpfull

forvalues nsim=1/1000 {
      *print i
      di "`nsim'"

      foreach b_yx of numlist 0 0.5 1 {
            foreach b_yu of numlist 0 0.5 1 {
                  foreach b_ru of numlist 0 0.5 1 {
                        foreach b_rz of numlist 0 0.5 1 {
                              clear
                              quietly set obs 1000
                              gen z=rnormal(0,1)
                              gen x=rnormal(0,1)
                              gen u=rnormal(0,1)
                              gen y=rnormal(`b_yx'*x + `b_yu'*u,1)
                              gen r=rnormal(`b_ru'*u + `b_rz'*z,1)

                              gen xmiss=x
                              quietly replace xmiss=. if r>0
                              quietly save `tmpfull', replace

                        *MI with Y
                        quietly mi set flong
                        quietly mi register imputed xmiss
                        quietly mi register regular y
                        quietly mi impute regress xmiss = y, add(5)
                        quietly mi estimate: regress y xmiss
                        local beta_x = e(b_mi)[1,1]
                        local se_x = sqrt(e(V_mi)[1,1])

                        *MI with X and Z
                        *restore simulated data i.e. before imputation
                        use `tmpfull', clear
                        quietly mi set flong
                        quietly mi register imputed xmiss
                        quietly mi register regular y z
                        quietly mi impute regress xmiss = y z, add(5)
```

```
                              quietly mi estimate: regress y xmiss

post `simloop' (`nsim') (`b_yx') (`b_yu') (`b_ru') (`b_rz')
(`beta_x') (`se_x') (e(b_mi)[1,1]) (sqrt(e(V_mi)[1,1]))
                         }
                    }
               }
          }
}
postclose `simloop'

* 3. Additional bias - Binary X
tempname simloop
postfile `simloop' int(nsim) float(b_yx b_yu b_ru b_rz beta_x_y
se_x_y beta_x_yz se_x_yz) using
"MIaddbias_illustration_scen2Xbin.dta", replace

*Create a temporary file for storing simulated data
tempfile tmpfull

forvalues nsim=1/1000 {
     di "`nsim'"

     foreach b_yx of numlist 0 0.5 1 {
          foreach b_yu of numlist 0 0.5 1 {
               foreach b_ru of numlist 0 0.5 1 {
                    foreach b_rz of numlist 0 0.5 1 {
                         clear
                         quietly set obs 1000
                         gen z=rnormal(0,1)
                         gen x=rbinomial(1,0.5)
                         gen u=rnormal(0,1)
                         gen y=rnormal(`b_yx'*x + `b_yu'*u,1)
                         gen r=rnormal(`b_ru'*u + `b_rz'*z,1)

                         gen xmiss=x
                         quietly replace xmiss=. if r>0
                         quietly save `tmpfull', replace

                         *MI with Y
                         quietly mi set flong
                         quietly mi register imputed xmiss
                         quietly mi register regular y
                         quietly mi impute logit xmiss = y, add(5)
                         quietly mi estimate: regress y xmiss
                         local beta_x = e(b_mi)[1,1]
                         local se_x = sqrt(e(V_mi)[1,1])

                         *MI with X and Z
                    *restore simulated data i.e. before imputation
                    use `tmpfull', clear
                    quietly mi set flong
                    quietly mi register imputed xmiss
                    quietly mi register regular y z
                    quietly mi impute logit xmiss = y z, add(5)
                    quietly mi estimate: regress y xmiss
```

```
post `simloop' (`nsim') (`b_yx') (`b_yu') (`b_ru') (`b_rz')
(`beta_x') (`se_x') (e(b_mi)[1,1]) (sqrt(e(V_mi)[1,1]))
                        }
                  }
            }
      }
}
postclose `simloop'

* 4. Additional bias - Binary Y
tempname simloop
postfile `simloop' int(nsim) float(b_yx b_yu b_ru b_rz beta_x_marg
se_x_marg beta_x_cra se_x_cra beta_x_miy se_x_miy beta_x_miyz
se_x_miyz) using "MIaddbias_illustration_scen2Ybin.dta", replace

*Create a temporary file for storing simulated data
tempfile tmpfull

forvalues nsim=1/1000 {
      di "`nsim'"

      foreach b_yx of numlist 0 0.5 1 {
            foreach b_yu of numlist 0 0.5 1 {
                  foreach b_ru of numlist 0 0.5 1 {
                        foreach b_rz of numlist 0 0.5 1 {
                              clear
                  quietly set obs 1000
                  gen z=rnormal(0,1)
                  gen x=rnormal(0,1)
                  gen u=rnormal(0,1)
                  gen y=rbinomial(1,invlogit(`b_yx'*x + `b_yu'*u))
                  gen r=rnormal(`b_ru'*u + `b_rz'*z,1)
                  gen ymiss=y
                  quietly replace ymiss=. if r>0
                  quietly save `tmpfull', replace

                              *Full data estimate
                              quietly logit y x
                              local beta_xmarg=e(b)[1,1]
                              local se_xmarg=sqrt(e(V)[1,1])

                              *CRA
                              quietly logit ymiss x
                              local beta_xcra=e(b)[1,1]
                              local se_xcra=sqrt(e(V)[1,1])

                              *MI with Y
                              quietly mi set flong
                              quietly mi register imputed ymiss
                              quietly mi register regular x
                              quietly mi impute logit ymiss = x, add(5)
                              quietly mi estimate: logit ymiss x
                              local beta_x = e(b_mi)[1,1]
                              local se_x = sqrt(e(V_mi)[1,1])
```

```
                              *MI with X and Z
                        *restore simulated data i.e. before imputation
                        use `tmpfull', clear
                        quietly mi set flong
                        quietly mi register imputed ymiss
                        quietly mi register regular x z
                        quietly mi impute logit ymiss = x z, add(5)
                        quietly mi estimate: logit ymiss x

post `simloop' (`nsim') (`b_yx') (`b_yu') (`b_ru') (`b_rz')
(`beta_xmarg') (`se_xmarg') (`beta_xcra') (`se_xcra') (`beta_x')
(`se_x') (e(b_mi)[1,1]) (sqrt(e(V_mi)[1,1]))
                        }
                  }
            }
      }
}
postclose `simloop'


*** Scenario 3  ***
*** Setting 1: Y partially observed ***
* 1. Verification of bias
*Define postfile to store results
tempname simloop
postfile `simloop' int(i) float(b_yx beta_yx beta_yx_cond_r
beta_yx_cond_rz theor_bias emp_bias theor_addbias emp_addbias) using
"sim_yxzr_scen3.dta", replace

forvalues i=1/1000 {
      *di "`i'"
      clear
      *SD
      local s_z=runiform(0,2)
      local s_r=runiform(0,2)
      local s_x=runiform(0,2)
      local s_y=runiform(0,2)
      *beta
      local b_yx=runiform(0,2)
      local b_ry=runiform(0,2)
      local b_rx=runiform(0,2)
      local b_rz=runiform(0,2)

      *RVs
      quietly set obs 100000
      gen z=rnormal(0,`s_z')
      gen x=rnormal(0,`s_x')
      gen y=rnormal(`b_yx'*x,`s_y')
      gen r=rnormal(`b_ry'*y + `b_rx'*x + `b_rz'*z,`s_r')

      *Estimate beta_YX
      quietly regress y x
      local beta_yx=e(b)[1,1]

      *Estimate beta_YX|R
      quietly regress y x r
```

```
      *Store estimate
      local beta_yx_cond_r=e(b)[1,1]

      *Estimate beta_YX|R,Z
      quietly regress y x r z
      *Store estimate
      local beta_yx_cond_rz=e(b)[1,1]

      *Calculate theoretical and empirical bias and bias amp
      local emp_bias = `beta_yx_cond_r' - `beta_yx'
      local emp_addbias = `beta_yx_cond_rz' - `beta_yx_cond_r'

      local theor_bias=-`b_yx'*`b_ry'*`s_y'^2*(`b_ry' +
(`b_rx'/`b_yx'))/(`b_ry'^2*`s_y'^2 + `s_r'^2 + `b_rz'^2*`s_z'^2)
      local theor_addbias=`b_yx'*`b_ry'*`s_y'^2*(`b_ry' +
(`b_rx'/`b_yx'))*((1/(`b_ry'^2*`s_y'^2 + `s_r'^2 +
`b_rz'^2*`s_z'^2)) - (1/(`b_ry'^2*`s_y'^2 + `s_r'^2)))

post `simloop' (`i') (`b_yx') (`beta_yx') (`beta_yx_cond_r')
(`beta_yx_cond_rz') (`theor_bias') (`emp_bias') (`theor_addbias')
(`emp_addbias')
}
postclose `simloop'


* 2. Illustration of max additional bias formula
*Define postfile to store results
postfile `simloop' float(b_yx b_ry b_rz b_rx max_addbias) using
"maxaddbias_illustration_scen3Ycts.dta", replace

foreach b_yx of numlist 0 0.25 0.5 0.75 1 {
      foreach b_ry of numlist 0 0.25 0.5 0.75 1 {
            foreach b_rz of numlist 0 0.25 0.5 0.75 1 {
                        foreach b_rx of numlist 0 0.25 0.5 0.75 1 {
                              clear
*Write in a different form to avoid division by zero
                              local max_addbias=(`b_yx'*`b_ry'^2 +
`b_ry'*`b_rx')*((1/(`b_ry'^2 + 1 + `b_rz'^2)) - (1/(`b_ry'^2 + 1)))

post `simloop' (`b_yx') (`b_ry') (`b_rz') (`b_rx') (`max_addbias')
                        }
            }
      }
}
postclose `simloop'

* 3. Binary Y
tempname simloop
postfile `simloop' int(nsim) float(b_yx b_ry b_rz b_rx beta_x_x
se_x_x beta_x_xz se_x_xz) using
"MIaddbias_illustration_scen3Ybin.dta", replace

*Create a temporary file for storing simulated data
tempfile tmpfull

forvalues nsim=1/1000 {
```

```
	di "`nsim'"

	foreach b_yx of numlist 0 0.5 1 {
		foreach b_ry of numlist 0 0.5 1 {
			foreach b_rz of numlist 0 0.5 1 {
				foreach b_rx of numlist 0 0.5 1 {
						clear
			quietly set obs 1000
			gen z=rnormal(0,1)
			gen x=rnormal(0,1)
			gen y=rbinomial(1,invlogit(`b_yx'*x ))
			gen r=rnormal(`b_ry'*y + `b_rx'*x + `b_rz'*z,1)

			gen ymiss=y
			quietly replace ymiss=. if r>0
			quietly save `tmpfull', replace

			*MI with X
			quietly mi set flong
			quietly mi register imputed ymiss
			quietly mi register regular x
			quietly mi impute logit ymiss = x, add(5)
			quietly mi estimate: logistic ymiss x
			local beta_x = e(b_mi)[1,1]
			local se_x = sqrt(e(V_mi)[1,1])

			*MI with X and Z
			*restore simulated data i.e. before imputation
			use `tmpfull', clear
			quietly mi set flong
			quietly mi register imputed ymiss
			quietly mi register regular x z
			quietly mi impute logit ymiss = x z, add(5)
			quietly mi estimate: logistic ymiss x

post `simloop' (`nsim') (`b_yx') (`b_ry') (`b_rz') (`b_rx')
(`beta_x') (`se_x') (e(b_mi)[1,1]) (sqrt(e(V_mi)[1,1]))
				}
			}
		}
	}
}
postclose `simloop'

*** Setting 2. X partially observed ***
* 1. Verification of maximum bias of the Y coefficient
*Define postfile to store results
tempname simloop
postfile `simloop' int(i) float(b_yx beta_xy beta_xy_cond_r
beta_xy_cond_rz theor_bias emp_bias theor_addbias emp_addbias) using
"sim_yxzr_scen3_impX.dta", replace

forvalues i=1/1000 {
	clear
	*SD
	local s_z=runiform(0,2)
```

```
	local s_r=runiform(0,2)
	local s_x=runiform(0,2)
	local s_y=runiform(0,2)
	*beta
	local b_yx=runiform(0,2)
	local b_ry=runiform(0,2)
	local b_rx=runiform(0,2)
	local b_rz=runiform(0,2)

	*RVs
	quietly set obs 100000
	gen z=rnormal(0,`s_z')
	gen x=rnormal(0,`s_x')
	gen y=rnormal(`b_yx'*x,`s_y')
	gen r=rnormal(`b_ry'*y + `b_rx'*x + `b_rz'*z,`s_r')

	*Estimate beta_XY
	quietly regress x y
	local beta_xy=e(b)[1,1]

	*Estimate beta_XY|R
	quietly regress x y r
	*Store estimate
	local beta_xy_cond_r=e(b)[1,1]

	*Estimate beta_XY|R,Z
	quietly regress x y r z
	*Store estimate
	local beta_xy_cond_rz=e(b)[1,1]

	*Calculate theoretical and empirical bias and bias amp
	local emp_bias = `beta_xy_cond_r' - `beta_xy'
	local emp_addbias = `beta_xy_cond_rz' - `beta_xy_cond_r'
	local b_xy=`b_yx'*`s_x'^2/(`b_yx'^2*`s_x'^2 + `s_y'^2)
	local theor_bias=-`b_xy'*`b_rx'*(`b_ry'*`s_y'^2/`b_yx' +
`b_rx'*`s_x'^2*(1 -
`b_yx'^2*`s_x'^2/(`b_yx'^2*`s_x'^2+`s_y'^2)))/(`b_rx'^2*`s_x'^2 +
`s_r'^2 + `b_rz'^2*`s_z'^2 -
(`b_yx'^2*`b_rx'^2*`s_x'^4/(`b_yx'^2*`s_x'^2+`s_y'^2)))
	local theor_addbias=`b_xy'*`b_rx'*((`b_ry'*`s_y'^2/`b_yx') +
`b_rx'*`s_x'^2*(1 - (`b_yx'^2*`s_x'^2/(`b_yx'^2*`s_x'^2+`s_y'^2))))*
///
		((1/(`b_rx'^2*`s_x'^2 + `s_r'^2 + `b_rz'^2*`s_z'^2 -
(`b_yx'^2*`b_rx'^2*`s_x'^4/(`b_yx'^2*`s_x'^2+`s_y'^2)))) -
(1/(`b_rx'^2*`s_x'^2 + `s_r'^2  -
(`b_yx'^2*`b_rx'^2*`s_x'^4/(`b_yx'^2*`s_x'^2+`s_y'^2)))))

post `simloop' (`i') (`b_xy') (`beta_xy') (`beta_xy_cond_r')
(`beta_xy_cond_rz') (`theor_bias') (`emp_bias') (`theor_addbias')
(`emp_addbias')
}
postclose `simloop'

* 2. Bias illustration
****** Varying the size of parameters - still use mu_z=0 and mu_u=0
and all error vars = 1*******
```

```
tempname simloop
postfile `simloop' int(nsim) float(b_yx b_ry b_rz b_rx beta_x_x
se_x_x beta_x_xz se_x_xz) using
"MIaddbias_illustration_scen3Xcts.dta", replace

*Create a temporary file for storing simulated data
tempfile tmpfull

forvalues nsim=1/1000 {
      *print i
      di "`nsim'"

      foreach b_yx of numlist 0 0.5 1 {
            foreach b_ry of numlist 0 0.5 1 {
                  foreach b_rz of numlist 0 0.5 1 {
                        foreach b_rx of numlist 0 0.5 1 {
                                    clear
                  quietly set obs 1000
                  gen z=rnormal(0,1)
                  gen x=rnormal(0,1)
                  gen y=rnormal(`b_yx'*x,1)
                  gen r=rnormal(`b_ry'*y + `b_rx'*x + `b_rz'*z,1)

                  gen xmiss=x
                  quietly replace xmiss=. if r>0
                  quietly save `tmpfull', replace

                  *MI with Y
                  quietly mi set flong
                  quietly mi register imputed xmiss
                  quietly mi register regular y
                  quietly mi impute regress xmiss = y, add(5)
                  quietly mi estimate: regress y xmiss
                  local beta_x = e(b_mi)[1,1]
                  local se_x = sqrt(e(V_mi)[1,1])

                  *MI with Y and Z
                  *restore simulated data i.e. before imputation
                  use `tmpfull', clear
                  quietly mi set flong
                  quietly mi register imputed xmiss
                  quietly mi register regular y z
                  quietly mi impute regress xmiss = y z, add(5)
                  quietly mi estimate: regress y xmiss

post `simloop' (`nsim') (`b_yx') (`b_ry') (`b_rz') (`b_rx')
(`beta_x') (`se_x') (e(b_mi)[1,1]) (sqrt(e(V_mi)[1,1]))
                        }
                  }
            }
      }
}
postclose `simloop'
```

```
* 3. Additional bias - Binary X
****** Varying the size of parameters - still use mu_z=0 and mu_u=0
and all error vars = 1*******

tempname simloop
postfile `simloop' int(nsim) float(b_yx b_ry b_rz b_rx beta_x_x
se_x_x beta_x_xz se_x_xz) using
"MIaddbias_illustration_scen3Xbin.dta", replace

*Create a temporary file for storing simulated data
tempfile tmpfull

forvalues nsim=1/1000 {
      *print i
      di "`nsim'"

      foreach b_yx of numlist 0 0.5 1 {
            foreach b_ry of numlist 0 0.5 1 {
                  foreach b_rz of numlist 0 0.5 1 {
                        foreach b_rx of numlist 0 0.5 1 {
                                    clear
            quietly set obs 1000
            gen z=rnormal(0,1)
            gen x=rbinomial(1,0.5)
            gen y=rnormal(`b_yx'*x,1)
            gen r=rnormal(`b_ry'*y + `b_rx'*x + `b_rz'*z,1)

            gen xmiss=x
            quietly replace xmiss=. if r>0
            quietly save `tmpfull', replace

            *MI with Y
            quietly mi set flong
            quietly mi register imputed xmiss
            quietly mi register regular y
            quietly mi impute logit xmiss = y, add(5)
            quietly mi estimate: regress y xmiss
            local beta_x = e(b_mi)[1,1]
            local se_x = sqrt(e(V_mi)[1,1])

            *MI with Y and Z
            *restore simulated data i.e. before imputation
            use `tmpfull', clear
            quietly mi set flong
            quietly mi register imputed xmiss
            quietly mi register regular y z
            quietly mi impute logit xmiss = y z, add(5)
            quietly mi estimate: regress y xmiss
post `simloop' (`nsim') (`b_yx') (`b_ry') (`b_rz') (`b_rx')
(`beta_x') (`se_x') (e(b_mi)[1,1]) (sqrt(e(V_mi)[1,1]))
                        }
                  }
            }
      }
}
postclose `simloop'
```

*Section S7. Stata code to perform the real data analysis*

```
* 1. Check mDAG associations
*IQ15 and maternal smoking
regress iq15 i.matsmoki i.bf_bin i.sex i.msoc_prof_nonman
i.mated_16plus i.parity_cat matage i.housing_cat

*Predictors of R_IQ15
logistic comp_caseiq15 i.matsmoki i.bf_bin i.sex i.msoc_prof_nonman
i.mated_16plus i.parity_cat matage i.housing_cat, coef

*2. CRA
regress iq15 i.bf_bin i.sex i.msoc_prof_nonman i.mated_16plus
i.parity_cat matage i.housing_cat

*3. MI using i.bfduration i.mated i.msoc to predict iq15 and vice
versa
quietly mi set flong
quietly mi register imputed iq15 bf_bin msoc_prof_nonman
mated_16plus parity_cat housing_cat
quietly mi register regular sex matage
mi impute chained (regress) iq15 (logit) bf_bin (ologit) parity_cat
(mlogit) housing_cat (logit) msoc_prof_nonman mated_16plus = i.sex
matage, ///
      add(100) burnin(20) /*dryrun*/ dots
mi estimate: regress iq15 i.bf_bin i.sex i.msoc_prof_nonman
i.mated_16plus i.parity_cat matage i.housing_cat

* 4. MI additionally using matsmok
use alspac_subv1, clear
count
*13923
quietly mi set flong
quietly mi register imputed iq15 bf_bin msoc_prof_nonman
mated_16plus parity_cat housing_cat matsmoki
quietly mi register regular sex matage
mi impute chained (regress) iq15 (logit) bf_bin (ologit) parity_cat
(mlogit) housing_cat (logit) msoc_prof_nonman mated_16plus matsmoki
= i.sex matage, ///
      add(100) burnin(20) /*dryrun*/ dots
mi estimate: regress iq15 i.bf_bin i.sex i.msoc_prof_nonman
i.mated_16plus i.parity_cat matage i.housing_cat

* 5. Map to theor bias
corr bf_bin matsmoki, covariance
gen var_X=r(Var_1)
gen var_Z=r(Var_2)

logistic comp_caseiq15 i.matsmoki i.bf_bin i.sex i.msoc_prof_nonman
i.mated_16plus i.parity_cat matage i.housing_cat, coef
gen logOR_Z=e(b)[1,2]
gen logOR_X=e(b)[1,4]

/* Substituting in all terms gives: */
gen maxbiasamp=1+((0.6*logOR_Z)^2*var_Z/(1-(0.6*logOR_Z)^2*var_Z-
(0.6*logOR_X)^2*var_X))
```
